# Life-course neighbourhood deprivation and brain structure in older adults: The Lothian Birth Cohort 1936

**DOI:** 10.1101/2023.04.13.23288523

**Authors:** Gergő Baranyi, Colin R. Buchanan, Eleanor L.S. Conole, Ellen V. Backhouse, Susana Muñoz Maniega, Maria Valdes Hernandez, Mark E. Bastin, Joanna Wardlaw, Ian J. Deary, Simon R. Cox, Jamie Pearce

## Abstract

Neighbourhood disadvantage may be associated with brain health but the importance at different stages of the life course is poorly understood. Utilizing the Lothian Birth Cohort 1936, we explored the relationship between residential neighbourhood deprivation from birth to late adulthood, and global and regional neuroimaging measures at age 73. We found that residing in disadvantaged neighbourhoods in mid- to late adulthood was associated with smaller total brain (*β*=-0.06; SE=0.02; *n*=390) and grey matter volume (*β*=-0.11; SE=0.03; *n*=390), thinner cortex (*β*=-0.15; SE=0.06; *n*=379), and lower general white matter fractional anisotropy (*β*=-0.19; SE=0.06; *n*=388). Regional analysis identified affected focal cortical areas and specific white matter tracts. Among individuals belonging to lower occupational social classes, the brain-neighbourhood associations were stronger, with the impact of neighbourhood deprivation accumulating across the life course. Our findings suggest that living in deprived neighbourhoods is associated with adverse brain morphologies, with occupational social class adding to the vulnerability.

## INTRODUCTION

In 2020, there were 727 million people aged 65 years and older globally, with their proportion in the world’s population (9.3%) expected to grow^1^. One out of six individuals will be over 65 by 2050^1^ and approximately 10% of this population will likely experience dementia (153 million) ^2^. Non-pathological cognitive decline heralds dementia and is much more common in older individuals: self-experienced decline in cognitive functioning affects one in four cognitively non-impaired adults over the age of 60^3^. Age-related changes in the brain structure can provide biologically traceable objective markers of cognitive function, revealing how the brain ages, and offering an early indicator of cognitive decline^4^. Accumulated evidence shows modest positive associations between cognitive function and total brain volume as well as various metrics of grey and white matter micro- and macrostructures^5^, and these relationships explain a significant proportion of individual differences in cognitive function and cognitive decline, especially in older ages^4, 6^.

Later-life cognitive function and decline has been linked to various neighbourhood-level characteristics^7, 8^, and there is some research connecting markers of brain morphology derived from magnetic resonance imaging (MRI) to physical and social features of the residential environment. Available cross-sectional studies suggest that neighbourhood deprivation is associated with structural brain differences among older adults, ^9–12^ which may partly explain the link between neighbourhood and cognitive function^10^. Moreover, residing in deprived areas may be a social stressor, which can amplify the impact of environmental threats on cognitive function^13^. Our understanding is more limited when it comes to the association between neighbourhood conditions and cortical disconnection^14^, the deterioration of the brain’s connective pathways, for example, by axonal demyelination^15^. In addition to volumetric changes, disruptions in the brain’s white matter also contribute to later-life cognitive decline^4, 15^. Diffusion MRI (dMRI) can quantify water molecular diffusion in white matter microstructure^16^. From the diffusion tensor model we can obtain commonly used indicators of white matter microstructure such as fractional anisotropy (i.e., directional coherence of water molecule diffusion), and mean diffusivity (i.e., magnitude of water molecule diffusion), which typically go down and up with age, respectively^15^.

From a methodological perspective, existing research on neighbourhood and brain health in late adulthood is limited by cross-sectional and short-term longitudinal study designs which are prone to bias and/or confounding. Fluid cognitive abilities start to decline after young adulthood^17^ and the prodromal phase of dementia can stretch over a decade until diagnosis^18^. It is plausible that declining cognitive function and/or preclinical cognitive impairment impacts moving to neighbourhoods with specific amenities leading to reverse causation^19^. In addition to the impact of health status on moving, there is a degree of continuity in people’s residential context across their life course, with neighbourhood disadvantage earlier in life predicting neighbourhood context later^7, 20^, increasing the likelihood of erroneously identifying later-life exposures as more relevant. Applying the life-course approach (i.e. examining the long-term impact of social and physical exposures on later health and diseases risk^21^) has the potential to overcome methodological biases, ^7^ and it has been applied to explore individual-level risk factors of brain health among older adults^22^. Still, reconstructing objectively measured historical neighbourhood context over several decades remains a challenge and is not often done^19, 23^.

The present life-course study investigates whether living in deprived neighbourhoods from birth onwards was associated with global and regional brain health among older adults. Neighbourhood deprivation was operationalised as objectively measured area-level social disadvantage and it was linked to the residential history of participants in the Lothian Birth Cohort 1936 (LBC1936). We tested four exposure models to explain individual differences in grey and white matter macro- and microstructure at age 73 years: exposure to neighbourhood deprivation in childhood (0-19 years), in young adulthood (20-39 years), in mid- to late adulthood (40-69 years), and accumulated neighbourhood deprivation across the life course (0-69 years). Analyses were fitted with linear regression within the structural equation modelling framework applying full information maximum likelihood estimation, which allowed optimal model estimation using all available data (see details in Methods). Little is known about whether some population groups show stronger associations between neighbourhood deprivation and brain health; therefore, we explored effect modification by sex, apolipoprotein E (*APOE*) ε4 allele status, and individual-level social disadvantage in childhood and adulthood.

## RESULTS

There were 689 individuals with at least one global brain measure available, the total sample size slightly varied across the eight global brain outcomes (Table 1). The average age was 72.68 years at the time of MRI acquisition, 52.69% of participants were male and 29.66% were *APOE* ε4 allele carriers. Individuals with socially advantaged family backgrounds remained more likely to be socially advantaged as adults (χ^2^= 30.42; p<0.001); 25.76% and 57.82% of the sample belonged to higher occupational social class in childhood and in adulthood, respectively (Table 1). Neighbourhood deprivation could be only linked to participants residing in Edinburgh: childhood, young adulthood, mid- to late adulthood, and accumulated neighbourhood deprivation scores were available for 316, 388, 400, and 285 participants, respectively. Average deprivation scores decreased across participants’ lives (Table 1), but they remained positively intercorrelated (*r*=0.26-0.57) (Supplementary Table 1). Individuals living in more deprived neighbourhoods throughout their life-course were more likely to belong to lower occupational social classes in childhood and adulthood (suggesting social segregation by geography and restricted social mobility), had lower childhood IQ scores, and spent fewer years in full-time education (Supplementary Table 2).

**Table 1:** Characteristics of study participants included in the analyses.

| Characteristics | Total number | Mean ± SD / number (%) |
| --- | --- | --- |
| Global brain measures, mean ± SD<sup>a</sup> | | |
| Total brain volume (cm<sup>3</sup>) | 658 | 988.97 ± 89.44 |
| Grey matter volume (cm<sup>3</sup>) | 658 | 471.56 ± 44.71 |
| Normal-appearing white matter volume (cm<sup>3</sup>) | 658 | 475.01 ± 50.66 |
| White matter hyperintensity volume (cm<sup>3</sup>) | 672 | 12.06 ± 12.84 |
| Surface area (cm<sup>2</sup>) | 636 | 1533.72 ± 144.92 |
| Mean cortical thickness (mm) | 636 | 2.26 ± 0.10 |
| General fractional anisotropy (standardized unit)<sup>b</sup> | 665 | 0 ± 1 |
| General mean diffusivity (standardized unit)<sup>b</sup> | 665 | 0 ± 1 |
| Neighbourhood deprivation, mean ± SD | | |
| In childhood (age 0-19 years) | 316 | 0.57 ± 3.39 |
| In young adulthood (age 20-39 years) | 388 | -0.85 ± 2.80 |
| In mid- to late adulthood (age 40-69 years) | 400 | -2.35 ± 2.74 |
| Accumulated (age 0-69 years) | 285 | -2.00 ± 6.84 |
| Intracranial volume (cm<sup>3</sup>), mean ± SD | 680 | 1450.83 ± 140.52 |
| Age in years, mean ± SD | 689 | 72.68 ± 0.73 |
| Sex, number (%) | 689 |  |
| Male |  | 363 (52.69%) |
| Female |  | 326 (47.31%) |
| Father's occupational social class, number (%) | 629 |  |
| High (professional-managerial) |  | 162 (25.76%) |
| Low (skilled, partly skilled, and unskilled) |  | 467 (74.24%) |
| APOE ε4 allele status, number (%) | 654 | |
| ε4 carriers | | 194 (29.66%) |
| Not ε4 carriers | | 460 (70.34%) |
| Childhood IQ, mean ± SD | 652 | 100.80 (15.30) |
| Years spent in education, mean ± SD | 689 | 10.80 (1.14) |
| Adult occupational social class, number (%) | 678 |  |
| High (professional-managerial) |  | 392 (57.82%) |
| Low (skilled, partly skilled, and unskilled) |  | 286 (42.18%) |
| Stroke, number (%)<sup>c</sup> | 684 | 93 (13.60%) |
| Smoking status, number (%) | 689 |  |
| Current smoker |  | 56 (8.13%) |
| Ex-smoker |  | 310 (44.99%) |
| Never smoked |  | 323 (46.88%) |
| BMI, mean ± SD | 689 | 27.92 ± 4.49 |
| Self-reported history of |  |  |
| Cardiovascular diseases, number (%) | 689 | 187 (27.14%) |
| Diabetes, number (%) | 689 | 75 (10.89%) |
| Hypertension, number (%) | 689 | 339 (49.20%) |
| Stroke, number (%) | 689 | 48 (6.97%) |

<sup>a</sup> The total number of outcome observations defined the sample size of the analyses; specific associations were based on the number of pairwise complete observations for the respective covariate-outcome pairs.
<sup>b</sup> Operationalised as latent variable; in this table we report predicted values (standardized unit).
<sup>c</sup> Identified from MRI scans by a consultant neuroradiologist.

### Global brain measures

In the age and sex, (and intracranial volume for macrostructural outcome measures) adjusted model (i.e., Model 1), mid- to late adulthood neighbourhood deprivation was negatively associated with total brain volume, grey matter volume, mean cortical thickness, and general fractional anisotropy (*p*-value corrected for false discovery rate[*p_FDR_*]<0.05) (Table 2). These associations remained significant (*p_FDR_*<0.05) after further controlling for relevant life-course confounders (i.e., Model 2): neighbourhood deprivation in mid- to late adulthood was linked to smaller total brain (standardized effect size [*β*]=-0.06; standard error [SE]=0.02; *p_FDR_*=0.01; sample size [*N*]=658; pairwise complete observations [*n*]=390) and smaller grey matter volumes (*β*=-0.11; SE=0.03; *p_FDR_*=0.003; *N*=658; *n*=390), thinner cortex (*β*=-0.15; SE=0.06; *p_FDR_*=0.02; *N*=636, *n*=379), and lower general fractional anisotropy (*β*=-0.19; SE=0.06; *p_FDR_*=0.005; *N*=665, *n*=388) (Table 2). Although there were some significant associations with young adulthood and accumulated neighbourhood deprivation, these did not survive FDR correction.

**Table 2:** Associations between life-course models of neighbourhood deprivation and global brain measures

|  | Model 1<sup>a</sup> |  |  |  | Model 2<sup>b</sup> |  |  |  |
| --- | --- | --- | --- | --- | --- | --- | --- | --- |
| | $\beta$ | SE | $p$ | $p_{FDR}$ | $\beta$ | SE | $p$ | $p_{FDR}$ |
| *Childhood neighbourhood deprivation (n=296 to 311)* |  |  |  |  |  |  |  |  |
| Total brain volume | -0.03 | 0.02 | 0.16 | 0.70 | -0.04 | 0.02 | 0.10 | 0.38 |
| Grey matter volume | -0.04 | 0.03 | 0.18 | 0.70 | -0.05 | 0.03 | 0.09 | 0.38 |
| Normal-appearing white matter volume | 0.01 | 0.03 | 0.76 | 0.87 | 0.02 | 0.03 | 0.61 | 0.81 |
| White matter hyperintensity volume | -0.01 | 0.05 | 0.81 | 0.87 | -0.03 | 0.05 | 0.58 | 0.81 |
| Cortical surface area | -0.01 | 0.03 | 0.68 | 0.87 | -0.01 | 0.03 | 0.84 | 0.95 |
| Mean cortical thickness | -0.06 | 0.06 | 0.33 | 0.78 | -0.07 | 0.06 | 0.25 | 0.56 |
| General fractional anisotropy<sup>c</sup> | -0.05 | 0.06 | 0.39 | 0.78 | -0.07 | 0.06 | 0.28 | 0.56 |
| General mean diffusivity<sup>c</sup> | -0.01 | 0.06 | 0.87 | 0.87 | 0.00 | 0.06 | 0.96 | 0.95 |
| *Young adulthood neighbourhood deprivation (n=367 to 383)* |  |  |  |  |  |  |  |  |
| Total brain volume | -0.01 | 0.02 | 0.60 | 0.69 | -0.02 | 0.02 | 0.36 | 0.41 |
| Grey matter volume | -0.03 | 0.03 | 0.26 | 0.38 | -0.05 | 0.03 | 0.12 | 0.22 |
| Normal-appearing white matter volume | 0.04 | 0.03 | 0.21 | 0.38 | 0.05 | 0.03 | 0.14 | 0.22 |
| White matter hyperintensity volume | -0.01 | 0.05 | 0.82 | 0.82 | -0.04 | 0.05 | 0.47 | 0.47 |
| Cortical surface area | -0.04 | 0.03 | 0.17 | 0.38 | -0.05 | 0.03 | 0.06 | 0.18 |
| Mean cortical thickness | -0.14 | 0.05 | 0.01 | 0.06 | -0.10 | 0.05 | 0.07 | 0.18 |
| General fractional anisotropy<sup>c</sup> | -0.12 | 0.06 | 0.04 | 0.15 | -0.14 | 0.06 | 0.02 | 0.15 |
| General mean diffusivity<sup>c</sup> | -0.06 | 0.06 | 0.28 | 0.38 | -0.06 | 0.06 | 0.35 | 0.41 |
| *Mid- to late adulthood neighbourhood deprivation (n=379 to 396)* |  |  |  |  |  |  |  |  |
| Total brain volume | **-0.05** | **0.02** | **0.010** | **0.02** | **-0.06** | **0.02** | **0.004** | **0.01** |
| Grey matter volume | **-0.09** | **0.03** | **0.003** | **0.01** | **-0.11** | **0.03** | **<0.001** | **0.003** |
| Normal-appearing white matter volume | -0.06 | 0.03 | 0.05 | 0.08 | -0.07 | 0.03 | 0.03 | 0.05 |
| White matter hyperintensity volume | 0.04 | 0.05 | 0.39 | 0.45 | 0.05 | 0.05 | 0.35 | 0.40 |
| Cortical surface area | -0.03 | 0.03 | 0.22 | 0.29 | -0.04 | 0.03 | 0.16 | 0.22 |
| Mean cortical thickness | **-0.19** | **0.05** | **<0.001** | **0.002** | **-0.15** | **0.06** | **0.01** | **0.02** |
| General fractional anisotropy<sup>c</sup> | **-0.14** | **0.05** | **0.008** | **0.02** | **-0.19** | **0.06** | **0.001** | **0.006** |
| General mean diffusivity<sup>c</sup> | -0.02 | 0.06 | 0.70 | 0.70 | 0.03 | 0.06 | 0.68 | 0.68 |
| *Accumulated neighbourhood deprivation (n=268 to 281)* |  |  |  |  |  |  |  |  |
| Total brain volume | -0.04 | 0.02 | 0.05 | 0.16 | -0.05 | 0.03 | 0.03 | 0.13 |
| Grey matter volume | -0.06 | 0.03 | 0.06 | 0.16 | -0.09 | 0.04 | 0.01 | 0.11 |
| Normal-appearing white matter volume | 0.00 | 0.03 | 0.98 | 0.98 | 0.02 | 0.04 | 0.69 | 0.69 |
| White matter hyperintensity volume | 0.00 | 0.05 | 0.93 | 0.98 | -0.03 | 0.06 | 0.62 | 0.69 |
| Cortical surface area | -0.03 | 0.03 | 0.34 | 0.46 | -0.03 | 0.04 | 0.50 | 0.67 |
| Mean cortical thickness | -0.16 | 0.06 | 0.01 | 0.09 | -0.11 | 0.07 | 0.13 | 0.27 |
| General fractional anisotropy<sup>c</sup> | -0.08 | 0.06 | 0.19 | 0.38 | -0.14 | 0.07 | 0.06 | 0.17 |
| General mean diffusivity<sup>c</sup> | -0.07 | 0.07 | 0.28 | 0.45 | -0.06 | 0.08 | 0.44 | 0.67 |

<sup>a</sup> Models were adjusted for sex and age (and intracranial volume for macrostructural measures).
<sup>b</sup> Models were adjusted for sex, age, (intracranial volume for macrostructural measures,) father's occupational social class and *APOE* $\epsilon 4$ allele status. In addition, young adulthood models were adjusted for childhood IQ and years spent in education, and mid- to late adulthood/ accumulation models also for adult occupational social class.
<sup>c</sup> No adjustment for intracranial volume.

### Local brain measures: vertex-wide analysis

We explored regional associations across the entire cortical surface using vertex-wise analysis. After applying Model 1 adjustment, greater young adulthood and greater mid- to late adulthood neighbourhood deprivation was associated with lower cortical volume, and a thinner cortex, while greater accumulated neighbourhood deprivation was associated with a thinner cortex (Supplementary Figure S1-3). After further controlling for relevant life-course confounders (Model 2) only mid- to late adulthood neighbourhood deprivation associations remained FDR-corrected significant (Supplementary Figure S4-6; Figure 1). Specifically, greater mid- to late adulthood neighbourhood deprivation (N=622; n=371) was associated with lower cortical volume (mean β=-0.05; β range: −0.27 to 0.14); smaller cortical surface area (mean β=-0.02; β range: −0.22 to 0.15); and a thinner cortex (mean β=-0.08; β range: −0.28 to 0.12). There was a spatial overlap in the areas identified across the three cortical measures, particularly between volume and area (Dice coefficient=0.33), and between volume and thickness (Dice coefficient=0.13). The significant regions were in the: posterior area of the left superior frontal gyrus (both volume and area); rostral area of the right middle frontal gyrus (volume); right parahippocampal cortex (volume); right isthmus cingulate cortex (volume); caudal area of the right middle temporal gyrus (volume and thickness); caudal area of the right inferior temporal gyrus (thickness); and a lateral area of the right superior parietal lobule (thickness) (Figure 1).

**Figure 1:**
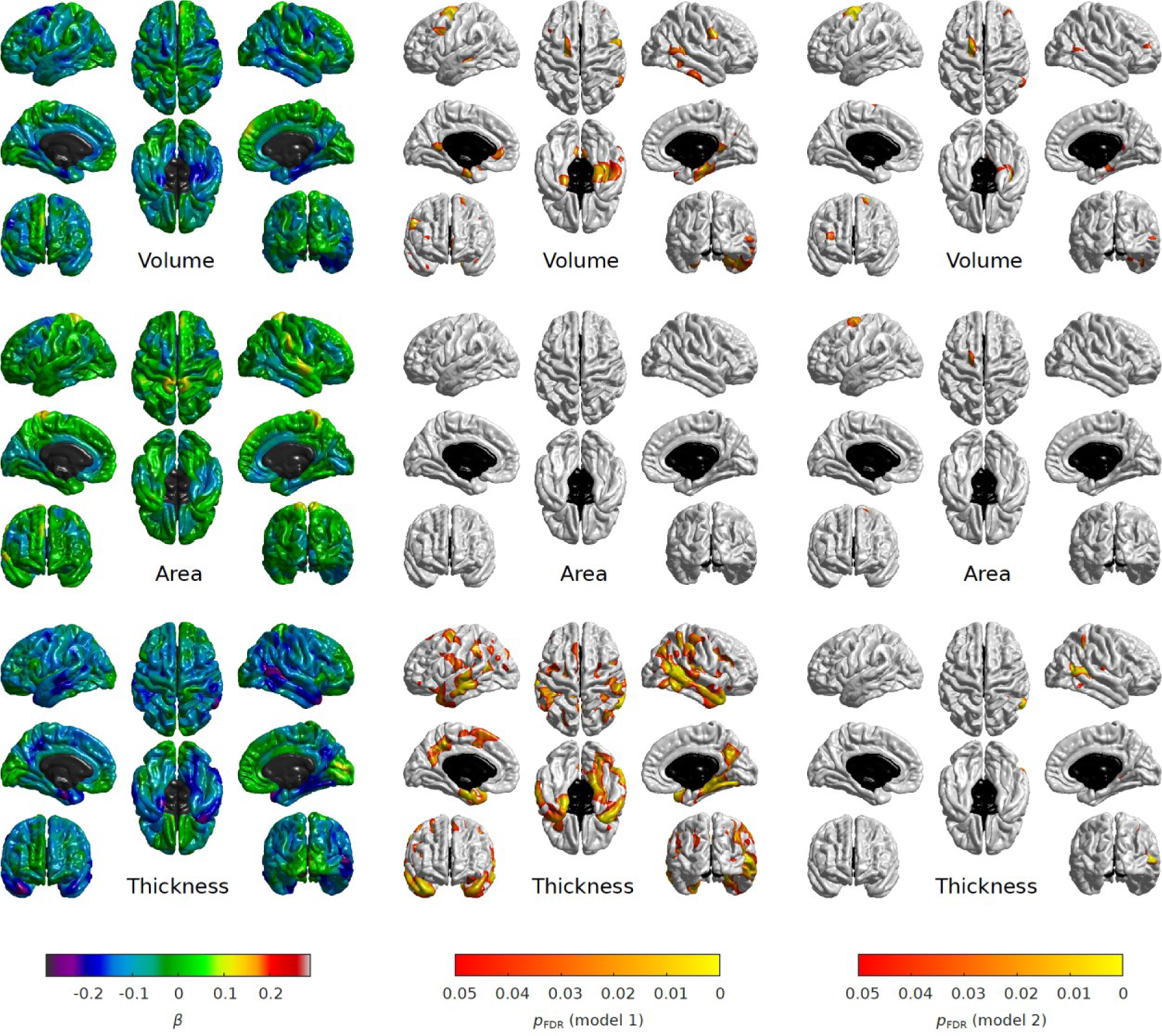
Regional associations between mid- to late adulthood neighbourhood deprivation and cortical properties (volume, surface area, and thickness). The fully adjusted standardized coefficients were obtained in linear regression models fitted within the structural equation modelling framework applying full information maximum likelihood estimation. Sample size was *N*=622, pairwise complete observations were n=371. The heatmaps show (left to right): standardised betas (Model 1), FDR-adjusted *p*-values for Model 1 (*p_FDR_*<0.05) and FDR-adjusted *p*-values for Model 2 (*p_FDR_*<0.05); the non-cortical mask is shown in black.

### Local brain measures: white matter tracts

Associations with neighbourhood deprivation were investigated in each of the twelve white matter tracts (Supplementary Figure 7). After FDR-correction, higher mid- to late adulthood neighbourhood deprivation was associated with lower fractional anisotropy in four tracts, in both Model 1 (Supplementary Table 5) and Model 2 (Supplementary Table 6): splenium of corpus callosum (Model 2: *β*=-0.19; SE=0.06; *p_FDR_*=0.003; *N*=652; *n*=379), right anterior thalamic radiation (Model 2: *β*=-0.19; SE=0.06; *p_FDR_*=0.003; *N*=662; *n*=371), right inferior longitudinal fasciculus (Model 2: *β*=-0.18; SE=0.06; *p_FDR_*=0.003; *N*=663; *n*=386), and left arcuate fasciculus (Model 2: *β*=-0.16; SE=0.06; *p_FDR_*=0.01; *N*=655; *n*=381). Right arcuate fasciculus became FDR-significant in Model 2 (*β*=-0.16; SE=0.06; *p_FDR_*=0.02; *N*=621; *n*=360) (Figure 2). Although there were no significant findings for mean diffusivity, stronger opposite direction associations in the same white matter tracts (e.g., splenium) reinforced findings for fractional anisotropy.

**Figure 2:**
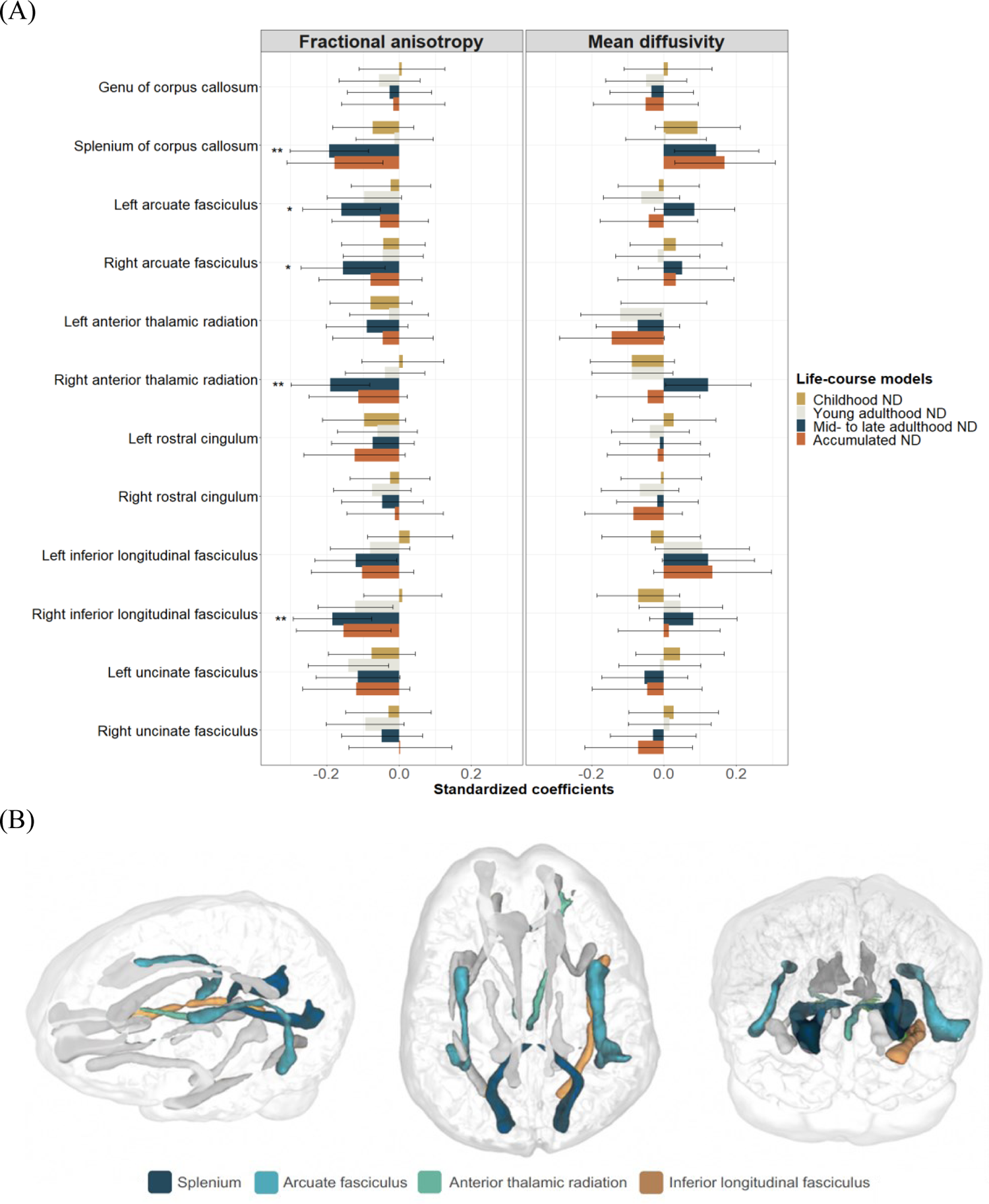
Association between life-course models of neighbourhood deprivation, and fractional anisotropy and mean diffusivity in twelve white matter tracts (A), and the location of false discovery rate (FDR)-adjusted significant tracts within the brain of a participant (B). Standardized coefficients with their 95% CI were obtained in linear regression models fitted within the structural equation modelling framework applying full information maximum likelihood estimation. Models were adjusted for sex, age, intracranial volume, father’s occupational social class, APOE ε4 allele status. In addition, young adulthood models were adjusted for childhood IQ and years spent in education, and mid- to late adulthood/ accumulation models also for adult occupational social class. Asterisks denote FDR-adjusted significance (\**p*_FDR_<0.05; \*\**p_FDR_*<0.01). ND=neighbourhood deprivation.

### Social class modifies neighbourhood-brain associations

We explored effect modification by age, *APOE* ε4 allele status, and occupational social class in childhood and adulthood. No FDR-adjusted differences were detected between females and males, or between ε4 carriers and non-ε4 carriers (Supplementary Table 7). The associations between neighbourhood deprivation and brain structural differences were stronger among individuals belonging to lower occupational social classes in childhood and in adulthood (i.e., skilled, partly skilled, and unskilled). Growing up in relatively disadvantaged households meant stronger negative association between young adulthood neighbourhood deprivation and late adulthood cortical surface area (*β*=-0.12; SE=0.04; *p_FDR_*=0.007; *N*=433; *n*=255) in comparison to higher social class households (i.e., professional-managerial).

Effect modification was found by own adult social class. Among individuals belonging to lower occupational social classes, higher childhood, young adulthood, and accumulated neighbourhood deprivation negatively associated with total brain and grey matter volumes, and – for childhood exposure – with general fractional anisotropy (Supplementary Table 7). When we estimated the associations of accumulated neighbourhood deprivation with total brain (*β*=-0.13; SE=0.04; *p_FDR_*=0.001; *N*=272; *n*=126) and grey matter volumes (*β*=-0.21; SE=0.06; *p_FDR_*=0.001; *N*=272; *n*=126) among socially disadvantaged individuals, these were 30.0% and 16.7% larger in magnitude, compared to any other life-course models (Supplementary Table 8). Figure 3 depicts associations between life-course models of neighbourhood deprivation and global brain measures by adult occupational social class.

**Figure 3:**
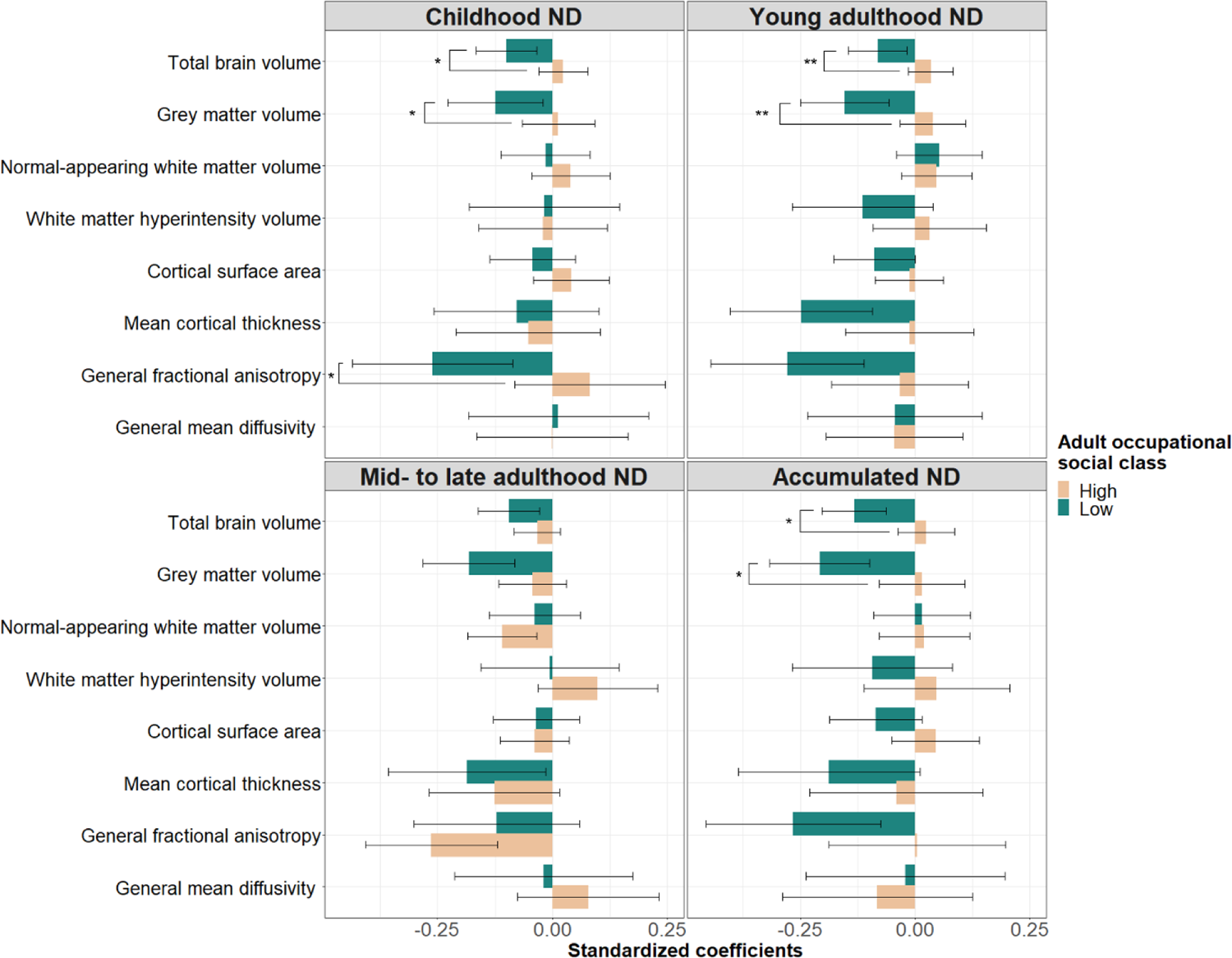
Association between life-course models of neighbourhood deprivation and global brain measures among participants belonging to high (i.e., professional-managerial) and low (i.e., skilled, partly skilled, and unskilled) occupational social classes in adulthood. Standardized coefficients with their 95% CI were obtained in linear regression models fitted within the structural equation modelling framework applying full information maximum likelihood estimation. Models were adjusted for sex, age, intracranial volume, father’s occupational social class, and APOE ε4 allele status; in addition, young adulthood/ mid- to late adulthood/ accumulation models were adjusted for childhood IQ and years spent in education. Asterisks denote FDR-adjusted significant interactions (\**p*_FDR_<0.05; \*\**p_FDR_*<0.01). ND=neighbourhood deprivation.

### Sensitivity analyses

After regressing young adulthood on childhood neighbourhood deprivation scores, effect sizes for the global brain measures dropped in magnitude with an average of 17%. After regressing mid- to late adulthood on young adulthood scores, associations were on average 15% lower; still, findings remained comparable (Supplementary Table 9). Adjusting for stroke identified from MRI scans did not affect the results (Supplementary Table 10). Controlling mid- to late adulthood exposure for health-related variables (i.e., BMI, smoking, self-reported medical diagnosis of chronic diseases) yielded 20% reduction of effect sizes on average, but findings remained comparable with the exemption of mean cortical thickness (−33%) (Supplementary Table 11). Applying a strict criterion with valid deprivation scores for each decade of the respective life-course model, reinforced the main findings. Importantly, it also suggested that accumulated neighbourhood deprivation significantly contributes to total brain volume (*β*=-0.09; SE=0.03; *p_FDR_*=0.03; *N*=658; *n*=222) with an effect size of 50% larger than for mid- to late adulthood exposure (Supplementary Table 12). When we excluded individuals from the sample with any signs of cognitive impairment, associations remained comparable with young adulthood exposure and general fractional anisotropy, as well as accumulated deprivation and total brain volume becoming FDR-significant (Supplementary Table 13). Finally, utilising linear regression with complete case analysis decreased power, but led to comparable nominal associations as the main analyses with general fractional anisotropy remaining (FDR-adjusted) significantly associated with neighbourhood deprivation in young adulthood and in mid-to-late adulthood (Supplementary Table 14).

## DISCUSSION

This study demonstrates that living in disadvantaged neighbourhoods across the life course is linked to brain structural differences among older adults. First, mid- to late adulthood neighbourhood deprivation was modestly associated with global and regional brain morphology. Participants living in deprived neighbourhoods had smaller total brain and grey matter volumes, thinner cortex, and lower general fractional anisotropy. Cortical differences were identified in both hemispheres, mainly in the left superior frontal gyrus and the right middle temporal gyrus. Smaller areas of significance were in right inferior frontal gyrus, right parahippocampal cortex, right cingulate cortex, right middle temporal gyrus, and the right superior parietal lobule. Regional associations for white matter tracts included lower fractional anisotropy in the splenium, in the arcuate fasciculus, in the right anterior thalamic radiation, and in the right inferior longitudinal fasciculus. Second, individual social disadvantage across the life-course amplified the association of neighbourhood deprivation with brain structural differences. Participants growing up in disadvantaged households showed additional detrimental impacts of neighbourhood deprivation in young adulthood, materialising as smaller cortical surface area. Participants belonging to lower occupational social classes in adulthood showed more accumulated impact of neighbourhood deprivation on total brain and grey matter volume. Sensitivity analyses reinforced the main associations with mid- to late adulthood neighbourhood deprivation, especially for grey matter volume and general fractional anisotropy.

The brain’s macro and micro-structure is constantly changing across the human life course. Total brain volume, for example, increases throughout childhood and adolescence, and after a plateau in young adulthood, a steady volume loss occurs with further acceleration after the age of 60 years^24, 25^. Neighbourhood deprivation may have long-term impacts on brain morphology, where aspects of social and environmental change may disrupt aspects of brain development during critical periods. Our study suggests that exposure to higher neighbourhood disadvantage in mid- to late adulthood (age 40-69), a period characterised with brain degeneration^24, 25^, was linked to brain structural differences (i.e., reduced total brain and grey matter volume, thinner cortex and lower general fractional anisotropy). Available cross-sectional evidence supports these findings^9–12, 14, 26^. Living in disadvantaged areas is associated with smaller total brain volume^9^, and with cortical thinning in Alzheimer’s disease signature regions^10^ in older age, but not with white matter hyperintensities^11^ – in line with our findings. Comparable associations were found in midlife for global brain outcomes^12^, for surface area and cortical thickness in regions linked to language and executive functions^26^, as well as for white matter fractional anisotropy^14^. The specific cortical regions we identified overlap with Alzheimer-relevant areas and are thought to be involved in executive function and perception (left superior frontal), emotion (right middle temporal and right isthmus cingulate), social cognition and working memory (right middle frontal), and imagination and perception (right superior parietal lobule); the identified white matter tracts are potentially involved in production and understanding of language (arcuate fasciculus), executive function (anterior thalamic radiation), and visual and semantic processing (inferior longitudinal fasciculus). ^27^ However, further studies are required to confirm regional associations.

Individual social disadvantage across the life course modified the relationship between neighbourhood deprivation and brain morphology, whereas there were no effect modifications by sex and *APOE* ε4 allele status (in line with previous studies on neighbourhood exposures^9, 28^). Interaction between family and neighbourhood disadvantage in childhood has been observed in cross-sectional studies; children from disadvantaged families living in more deprived areas have lower cortical thickness^29^, and stronger age-related decrease in cortical thickness^30^ in comparison to children from advantaged families. Increasing evidence shows that material disadvantage not only associates with brain structure during later-life, but also during childhood and adolescence^31^, with early life factors having long-term consequences likely lasting throughout life^22, 32^. Cross-sectional studies found smaller global and regional cortical surface area^33^, smaller cortical^34^ and subcortical volumes^33^, and lower cortical thickness^29, 30^ among children exposed to greater neighbourhood deprivation. Social disadvantage (including area deprivation) also contributes to lower white matter integrity among newborns^35, 36^ and children^37^. Importantly, childhood neighbourhood deprivation can have long-term consequences for brain development with longitudinal studies showing lower white matter quantitative anisotropy^38^ and greater amygdala reactivity^39^ in younger adulthood. Our study adds a life course perspective to these findings, but also extends previous LBC1936 analysis finding null associations between father’s occupational social class and total brain volume^22^: multi-faced disadvantage (individual and area-level) in early life was associated with cortical surface area differences among older adults.

The more time individuals spent living in deprived neighbourhoods across their lives, the larger were the association between neighbourhood deprivation and brain structural differences; these findings were particularly marked for disadvantaged adults. Possibly, through the accumulation model, we may have captured area-level processes affecting the brain’s morphology both during its development in childhood and during its degeneration in mid- to late adulthood, and individuals with lower personal resources across the life course, including cognitive reserve^40^, were more vulnerable. Alternatively, it is also plausible that in our study accumulated neighbourhood deprivation only affected the brain during its degeneration phase (e.g., through cumulative allostatic load^41^), with the lagged impact of early life neighbourhood deprivation ‘kicking in’ in later adulthood. However, in the absence of neuroimaging data and area-level deprivation measured across the entire life course within the same individuals, these hypotheses cannot be verified and only present associations without causal implications. As such data are likely many years away from being feasible, future studies should apply pseudo-/ accelerated longitudinal designs in different age groups to further explore the reported interactions.

Residing in disadvantaged areas might influence brain morphology through multiple interrelated ways and the underlying processes may differ across the life course. Socioeconomically disadvantaged areas are likely to suffer from poorer housing conditions^42^, higher crime and violence^43^, have lower provision of high quality green space^44^, and residents are more likely to be exposed to higher levels of contaminants (e.g. air pollution^45^), poorer diet, and to health-damaging commodities and services, such as alcohol, fast food and tobacco outlets^46^; these, in turn, can associate with brain morphology and function (e.g. green space^28^, air pollution^47^). In childhood, the quality of local schools, in late adulthood the availability of neighbourhood resources and amenities (e.g., community and cultural centres), may be pertinent. Neighbourhood disadvantage may contribute to brain structural differences through multiple interrelated pathways. Research shows that children exposed to stressors in their social environment experience activated hypothalamic-pituitary-adrenal axis which leads to long-term dysregulation and changes in the brain^31, 39^. Investigated pathways in adulthood include inflammatory^14, 26^, neuroendocrine^12^ and cardiovascular^9^ mechanisms.

We describe associations between neighbourhood deprivation from birth to late adulthood and global and regional brain differences. Study strengths included the narrow age-range and homogeneity of participants, the availability of residential history and objectively measured neighbourhood disadvantage covering an exceptionally long period of ∼70 years, the detailed assessment of global and regional brain measures, and the availability of key life-course social, biological, and psychological confounders (e.g., father’s social class, *APOE* ε4 allele status, childhood IQ). There are limitations to acknowledge. First, our sample consisted of a relatively healthy, educated and urbanised group of older adults, with higher childhood IQ^48^ and lower risk of mortality^49^ in comparison to the population average, leading to selection and survival bias. Second, given the challenges of collecting objective neighbourhood data across the life course (i.e., deprivation scores could be obtained only for individuals residing in Edinburgh) our effective sample was moderate (ranging from 268 to 396), and it differed across the four life-course models. It is plausible that analyses for specific models were relatively underpowered in comparison to others; still, restricting the sample size to participants with observations available for all four models would have led to significant drop in relevant observations (>30%), increased type II error, and sample bias. Third, reconstructing neighbourhood deprivation indices across the 20^th^ century was challenged by the inconsistent availability of historical data and their varying spatial aggregation in official records. Although 1961 ward geographies likely do not align with participant’s self-defined neighbourhoods, they were necessary to provide a common spatial resolution to handle missing data^23^. Fourth, information on health status prior to late adulthood is not available in LBC1936, which likely increases the risk of unmeasured confounding through selective residential mobility. Fifth, MRI data was only available in late adulthood; therefore, we cannot ascertain whether there is a direct pathway between childhood area disadvantage and later-life brain outcomes, or childhood exposure first influenced childhood brain development. Last, we tested a large number of associations and, to reduce type 1 error rate, we highlighted only FDR-adjusted significant results. It is plausible that we missed important associations.

Future studies could usefully replicate our findings in larger and more socially and ethnically diverse samples with repeatedly measured neighbourhood exposures and brain outcomes across the life course, and with specific focus on downward and upward social and geographic mobility. Furthermore, testing life-course associations for subcortical structures, identifying specific neighbourhood features most likely contributing to brain health, systematically assessing causal pathways over the lifespan (e.g., hypothalamic-pituitary-adrenal axis, inflammation, cardiovascular risks), and understanding the double burden of disadvantaged people living in disadvantaged areas requires further attention.

## Conclusions

Living in deprived neighbourhoods across the life course was associated with worse brain health among older adults, which can be measured over and above individual-level differences. Neighbourhood deprivation during later-life (age 40-69 years) was associated with brain structural differences in older age, but we also found some evidence for the accumulating impact of neighbourhood deprivation across the entire life course, especially among socially disadvantaged individuals. The life course approach can provide useful insights into how the social environment might ‘get under the skin,’ and future research should apply it more often to understand differences and changes in brain morphology and related cognition. Greater understanding of relevant brain regions, social and physical neighbourhood features pertinent to brain health, and potential causal pathways require further research attention.

## METHODS

### Study participants

Data were drawn from the LBC1936, a longitudinal study of relatively healthy older adults born in 1936. LBC1936 was designed to follow up some participants of the Scottish Mental Survey 1947 – a nationwide general cognitive testing exercise carried out among all 1936-born children attending Scottish schools on June 4^th^, 1947 – with the main aim to study non-pathological cognitive ageing in later life^50, 51^. Surviving participants of the Scottish Mental Survey 1947 living in the City of Edinburgh and the surrounding Lothian region of Scotland were traced and contacted by the Lothian Health Board^51^. Between 2004 and 2007 (Wave 1; mean age=70 years), 1091 subjects underwent detailed social, cognitive and health assessments; since Wave 2 (2007-2010; mean age=73 years; *n*=866), neuroimaging data are also available^51^. In 2014 (mean age=78 years), a ‘life grid’ questionnaire was administered among surviving LBC1936 participants to collect retrospective account of their residential history for every decades from birth to date of completion^50^. Flashbulb memory prompts (e.g. 9/11 attacks in New York) and participant supplied personal events assisted recall^23^. 593 cohort members provided life-course addresses (84% completion rate) which were geocoded using automatic geocoders and historical building databases^23^.

### Neighbourhood deprivation

Life-course neighbourhood deprivation was operationalised as small area-level social disadvantage, computed for the City of Edinburgh once for every decades of the study with data derived from administrative records^23^. In 1941, 1951, 1961 and 1971, information on overcrowding, population density, infant mortality, tenure and amenities contributed to an index of multiple deprivation^23^; for 1981, 1991 and 2001, we utilised the Carstairs index of deprivation (i.e. male unemployment, overcrowding, car ownership, low social class) ^52^. Geographic data were aggregated to a common spatial resolution (1961 ward-level; *n*=23) in order to support estimating of missing indicators; *z*-scores were calculated to ensure comparability across the two indices^23^. We linked residential addresses to deprivation scores using 10-year intervals (e.g., 1941 score linked to 1936-1945 addresses). Exposures closer in time were highly correlated (*r*>0.61) (Supplementary Figure 8). Finally, three periods captured average exposures to neighbourhood deprivation during childhood (1936-1955), young adulthood (1956-1975) and mid- to late adulthood (1976-2005) for participants having any valid exposure data during these epochs^53^. Accumulated neighbourhood disadvantage was calculated as the mean exposure across the three periods, requiring valid measurement of neighbourhood deprivation (i.e., living in the City of Edinburgh) at least once for each.

### MRI acquisition

All brain data were acquired between 2007 and 2010 (Wave 2); the MRI acquisition parameters have been described previously^54^. All participants underwent brain MRI on a 1.5 T GE Signa Horizon HDx clinical scanner (General Electric, Milwaukee, WI) with a manufacturer supplied 8-channel phased-array head coil. High resolution 3D T_1_-weighted inversion-recovery prepared, fast spoiled gradient-echo volumes were acquired in the coronal plane with 160 contiguous 1.3 mm thick slices resulting in voxel dimensions of 1 × 1 × 1.3 mm. T_2_-weighted fast spin echo volumes were acquired in the coronal plane with 80 contiguous 2 mm thick slices resulting in voxel dimensions of 1 × 1 × 2 mm. For the dMRI protocol, single-shot spin-echo echo-planar (EP) diffusion-weighted whole-brain volumes (*b*=1000 s mm^−2^) were acquired in 64 noncollinear directions, along with seven T_2_-weighted volumes (*b*=0 s mm^−2^). Seventy-two contiguous axial 2 mm thick slices were acquired resulting in 2 mm isotropic voxels.

### Image processing

We assessed both global and regional brain measures. From the T_1_- and T_2_-weighted data, various tissue volumes were estimated as described previously^54^. Total brain volume was estimated as intracranial volume minus cerebrospinal fluid, and grey matter volume as total brain volume minus white matter volume. White matter volume was segmented into normal appearing-white matter and white matter hyperintensities, the latter defined as hyperintense areas (>3 mm in diameter) in white matter. Additionally, cortical reconstruction was performed with the FreeSurfer image analysis suite (http://surfer.nmr.mgh.harvard.edu) v5.1.0. Cortical surface analyses were then performed using the SurfStat MATLAB toolbox (http://www.math.mcgill.ca/keith/surfstat). Surfaces were aligned vertex-wise into a common space (the FreeSurfer average template) and spatially smoothed at 20 mm full width at half maximum, allowing sample-wide analyses of volume, area, and thickness across the cortex.

All raw dMRI data were converted from DICOM to NIfTI-1 format using TractoR v2.6.2^55^. Using tools freely available in the FSL toolkit v4.1.9 (FMRIB, Oxford University: http://www.fmrib.ox.ac.uk)^56^, data underwent brain extraction^57^ performed on the T_2_-weighted EP volumes acquired along with the dMRI data. The brain mask was applied to all volumes after correcting for systematic eddy-current induced imaging distortions and bulk patient motion using affine registration to the first T_2_-weighted EP volume of each participant^58^. For all dMRI volumes, diffusion tensors were fitted at each voxel and water diffusion measures were estimated for mean diffusivity and fractional anisotropy at each voxel. Tractography was performed using an established probabilistic algorithm with a two-fibre model per voxel (BEDPOSTX/ProbtrackX) ^59, 60^. Analysis of twelve major white matter tracts was performed using probabilistic neighbourhood tractography^55^. These tracts were the genu and splenium of the corpus callosum, left and right arcuate fasciculi, left and right anterior thalamic radiation, left and right rostral cingulum, left and right inferior longitudinal fasciculus, and left and right uncinate fasciculus (see Supplementary Figure 7 for their locations). All tracts were visually quality checked, and exclusions were made on a tract basis. Tract-averaged diffusion parameters (i.e., fractional anisotropy, mean diffusivity) weighted by the streamline visitation count were then calculated from all voxels by tract^54, 61^.

#### Covariates

Relevant confounders were selected based on the literature^7, 9^ and are presented in a directed acyclic graph (Supplementary Figure 9). Age and sex (female, male) were included in all presented models. *APOE* ε4 allele status (ε4 carriers, not ε4 carriers) is a genetic risk factor of cognitive decline^62^. Father’s occupational social class was classified into high (I/II: professional-managerial) and low classes (III/IV/V: skilled, partly skilled and unskilled) ^63^; the same categorisation was used for own social class in adulthood (for women, husband’s class was taken if higher). Childhood IQ was measured at age 11 years with the Moray House Test No 12^50^, and education was captured as years spent in full-time education. In sensitivity analyses, we considered a range of health-related variables, which can be theorised as confounders, but also as mediators between neighbourhood deprivation and brain health^7^. They were collected at the time of MRI acquisition (i.e., Wave 2) and included stroke identified from MRI scans by a consultant neuroradiologist (yes, no), body mass index (BMI), smoking status (current smoker, past smoker, never smoked), and self-reported medical diagnosis (yes, no) of stroke, diabetes, hypertension, and cardiovascular disease.

### Statistical analysis

Models were fitted with full-information maximum likelihood (FIML) estimation within structural equation modelling (SEM) using the *lavaan* package^64^ v0.6-12 in R v4.2.1^65^. FIML regression has the advantage of estimating model parameters based on all available information, including participant with missing variables, increasing power, and thus lowering type II error. Importantly, fitting models in the context of all available data for confounders enables to calculate model residuals in a larger and more comprehensive sample and to estimate the impact of exposure more accurately. FIML regression produces equivalent results to models handling missing data with multiple imputation^66^. We reported total sample size for each analysis (*N*) and the number of pairwise complete observations (*n*) for associations of interest. Goodness of fit indices were not provided for fully saturated models.

The primary analysis tested the associations between neighbourhood deprivation scores and global brain measures, including six macro-structural outcomes (total brain, grey matter, normal-appearing white matter, and white matter hyperintensity [log-transformed to approximate normal distribution^61^] volumes, cortical surface area, and mean cortical thickness) and two markers of white matter microstructure. Consistent with prior work^67^, general factors of fractional anisotropy and mean diffusivity across the twelve white matter tracts were estimated as latent factors; we included residual correlations between the splenium and the genu of corpus callosum, and between right and left sides of the bilateral tracts (see Supplementary Table 15 for fit indices and factor loadings). We provided *p_FDR_* correcting for multiple comparison^68^ between the eight global brain outcomes.

Local brain associations were explored in the secondary analyses. We estimated regional associations with life-course neighbourhood deprivation across the entire cortical surface. Vertex-wise analysis was performed in a common space (the FreeSurfer average template) for 327,684 cortical vertices using all MRI participants with cortical surface data. Three vertex-wise brain measures were assessed: cortical volume, surface area and thickness. Vertex measures were exported from SurfStat and then SEM with FIML was used to iteratively estimate standardised coefficients and the corresponding *p*-values by vertex for each neighbourhood deprivation exposure. Correction for multiple comparison was performed by FDR and the findings were presented in cortical surface maps. The spatial overlap between significant cortical regions was assessed by the Dice coefficient. ^69^ In addition to the general factors, we presented associations in each of the twelve white matter tracts after FDR correction.

We considered two sets of adjustment. Model 1 included age and sex; macro-structural global measures were also corrected for the premorbid brain size by adjusting for intracranial volume. Model 2 additionally adjusted for confounders picked individually for each life-course model using a directed acyclic graph (Supplementary Figure 9). Father’s social class and *APOE* ε4 allele status were confounders for all life-course models, childhood IQ and education for young adulthood, mid- to late adulthood and accumulation models, while adult occupational social class for mid- to late adulthood and accumulation models. Analyses for local and regional global brain measures were performed with both Model 1 and Model 2 adjustments, exploratory and sensitivity analysis were estimated using Model 2 adjustments (if applicable).

We performed exploratory analyses to test whether the associations between neighbourhood deprivation and global brain structure differed by sex, *APOE* ε4 allele status, father’s social class, and own adult social class. Models were fitted with interaction terms; population groups with FDR-significant differences were carried forward to multi-group SEM analysis. Six sensitivity analyses tested the robustness of associations with global brain measures. First, to address high correlation between neighbourhood deprivation scores across the three epochs, we regressed young adulthood on childhood scores, mid- to late adulthood on young adulthood scores within SEM by preserving their temporal ordering (S1). Second, we adjusted all models for stroke identified from MRI scans (S2). Third, mid- to late adulthood models were further adjusted for health-related variables (i.e., BMI, smoking status, self-reported medical diagnoses) (S3). For sensitivity analysis 2 and 3, we presented % change in standardized coefficients to aid comparison with the main model (i.e., Model 2). Fourth, we applied a strict criterion for computing life-course exposures: participants were required to have valid neighbourhood deprivation scores (i.e., live in the City of Edinburgh) for each decade within a sensitive period, and for each decade during their lives for the accumulated deprivation model (S4). Fifth, to reduce bias from recall inaccuracy during residential history recollection we excluded participants with cognitive impairment^53^, identified as either reporting dementia or scoring <24 at the Mini-Mental State Examination in any of the available LBC1936 follow-up waves (Waves 1-5) (S5). Sixth, we presented main analysis with traditionally performed linear regression with complete cases (S6).

## Data Availability

The LBCs’ study data have been the subject of many internal (within the University of Edinburgh) and external collaborations, which are encouraged. Those who have interests in outcomes other than cognitive domains are particularly encouraged to collaborate. Both LBC studies have clear data dictionaries which help researchers to discern whether the variables they wish to use are present; these provide a simple short title for each variable, alongside a longer, common-sense description/provenance of each variable. This information is available on the study website (https://www.ed.ac.uk/lothian-birth-cohorts) alongside comprehensive data grids listing all variables collected throughout both LBC studies and the wave at which they were introduced, an ‘LBC Data Request Form’ and example Data Transfer Agreement. Initially, the Data Request Form is e-mailed to the Lothian Birth Cohorts Director Dr Simon R. Cox for approval (via a panel comprising study co-investigators). Instances where approved projects require transfer of data or materials outside the University of Edinburgh require a formal Data Transfer Agreement or Material Transfer Agreement to be established with the host institution. The process is facilitated by a full-time LBC database manager – there is no charge.

For the purpose of open access, the author has applied a Creative Commons Attribution (CC BY) licence to any Author Accepted Manuscript version arising from this submission.

## ACKNOWLEDGEMENTS

The LBC1936 study was conducted according to the Declaration of Helsinki guidelines with ethical permission obtained from the Multi-Centre Research Ethics Committee for Scotland (MREC/01/0/56), Lothian Research Ethics Committee (Wave 1, LREC/2003/2/29), and the Scotland A Research Ethics Committee (Waves 2-4, 07/MRE00/58). Written consent was obtained from all participants.

## AUTHOR CONTRIBUTIONS

IJD and SRC obtained and managed the core LBC1936 dataset, JP obtained and processed the geographic data, and SMM, MVH, EB, MB, JW, IJD and SRC conducted the MRI acquisition and image processing. GB, IJD, SRC and JP conceived and designed the study. GB performed the main statistical analyses, CRB the vertex-wise analysis. ELSC prepared the glass brain plots (i.e., Figure 2B, Supplementary Figure 8). GB led the manuscript preparations, drafting and revisions. All authors participated in the interpretation of the findings, critically revised the manuscript, and approved the final version.

## Supplementary Material

**Supplementary Table 1:**
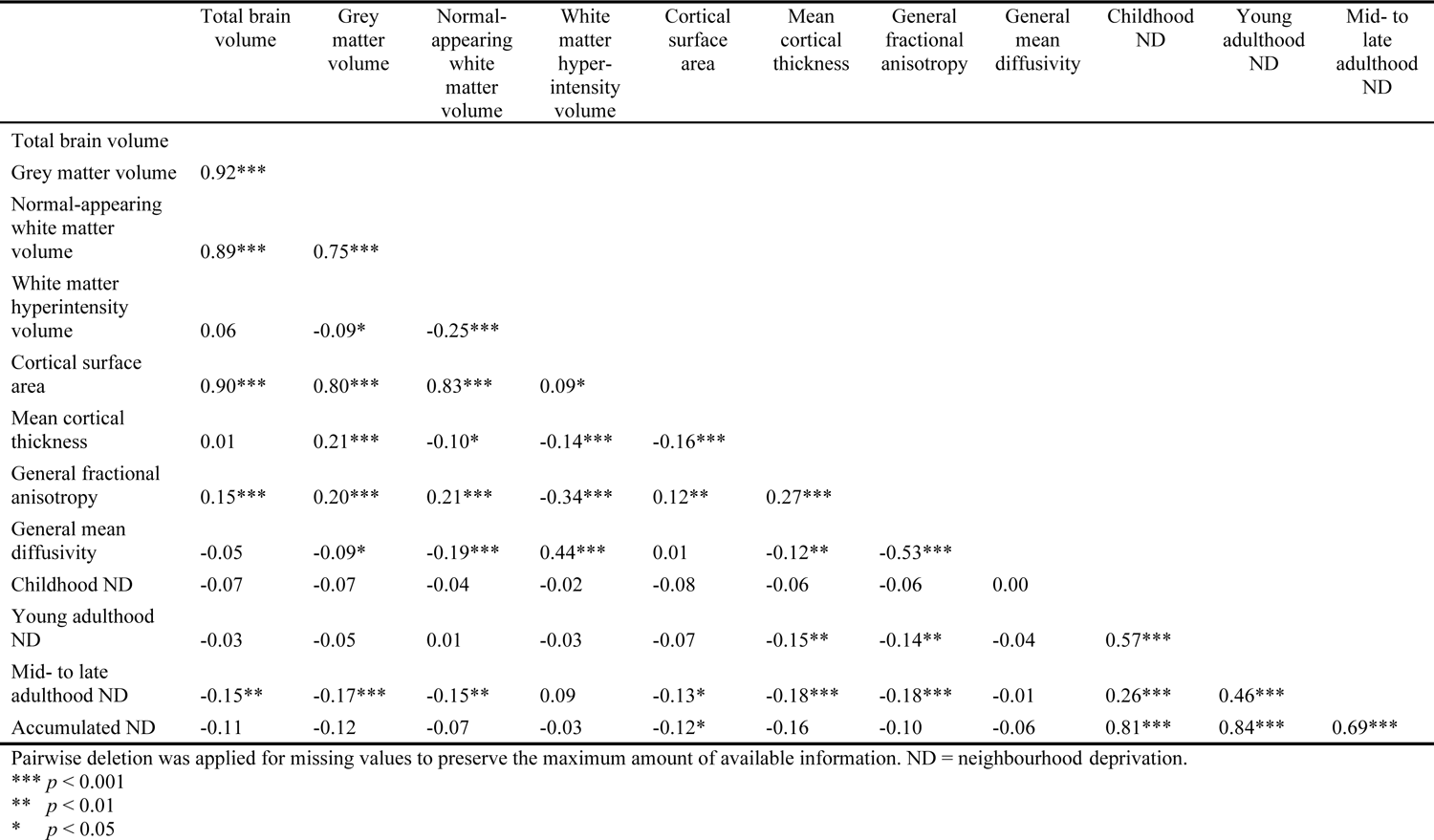
Pearson’s correlation coefficients between neighbourhood deprivation scores and global brain outcomes

**Supplementary Table 2:** Relation of main covariates to neighbourhood deprivation scores

|  | Childhood ND |  | Young adulthood ND |  | Mid- to late adulthood ND |  | Accumulated ND |  |
| --- | --- | --- | --- | --- | --- | --- | --- | --- |
|  | Mean (SD) | *p* | Mean (SD) | *p* | Mean (SD) | *p* | Mean (SD) | *p* |
| Sex |  |  |  |  |  |  |  |  |
| Male | 0.69 (3.51) |  | -0.84 (2.90) |  | -2.50 (2.81) |  | -2.01 (7.19) |  |
| Female | 0.44 (3.27) |  | -0.87 (2.69) |  | -2.19 (2.65) |  | -1.99 (6.47) |  |
| Father's occupational social class |  |  |  |  |  |  |  |  |
| High (professional-managerial) | -0.80 (3.38) | *** | -1.79 (2.72) | *** | -3.28 (2.40) | *** | -5.44 (6.15) | *** |
| Low (skilled, partly skilled and unskilled) | 1.06 (3.32) |  | -0.54 (2.74) |  | -2.06 (2.76) |  | -0.93 (6.68) |  |
| *APOE* ε4 allele status |  |  |  |  |  |  |  |  |
| ε4 carriers | 1.32 (3.25) | * | -0.56 (2.88) |  | -2.15 (2.90) |  | 0.01 (6.44) | *** |
| Not ε4 carriers | 0.23 (3.39) |  | -1.03 (2.74) |  | -2.50 (2.61) |  | -2.96 (6.76) |  |
| Adult occupational social class |  |  |  |  |  |  |  |  |
| High (professional-managerial) | -0.13 (3.29) | *** | -1.32 (2.66) | *** | -2.90 (2.56) | *** | -3.98 (6.08) | *** |
| Low (skilled, partly skilled and unskilled) | 1.48 (3.33) |  | -0.15 (2.87) |  | -1.47 (2.79) |  | 0.38 (6.97) |  |
|  | Pearson's *r* | *p* | Pearson's *r* | *p* | Pearson's *r* | *p* | Pearson's *r* | *p* |
| Childhood IQ | -0.18 | ** | -0.20 | *** | -0.23 | *** | -0.25 | *** |
| Years spent in education | -0.27 | *** | -0.28 | *** | -0.36 | *** | -0.42 | *** |

Pairwise deletion was applied for missing values to preserve the maximum amount of available information. Statistical analyses were based on two-sample t-tests for mean differences, and Pearson's correlation for associations between continuous variables. ND = neighbourhood deprivation.
\*\*\* $p < 0.001$
\*\* $p < 0.01$
\* $p < 0.05$

**Supplementary Table 3:**
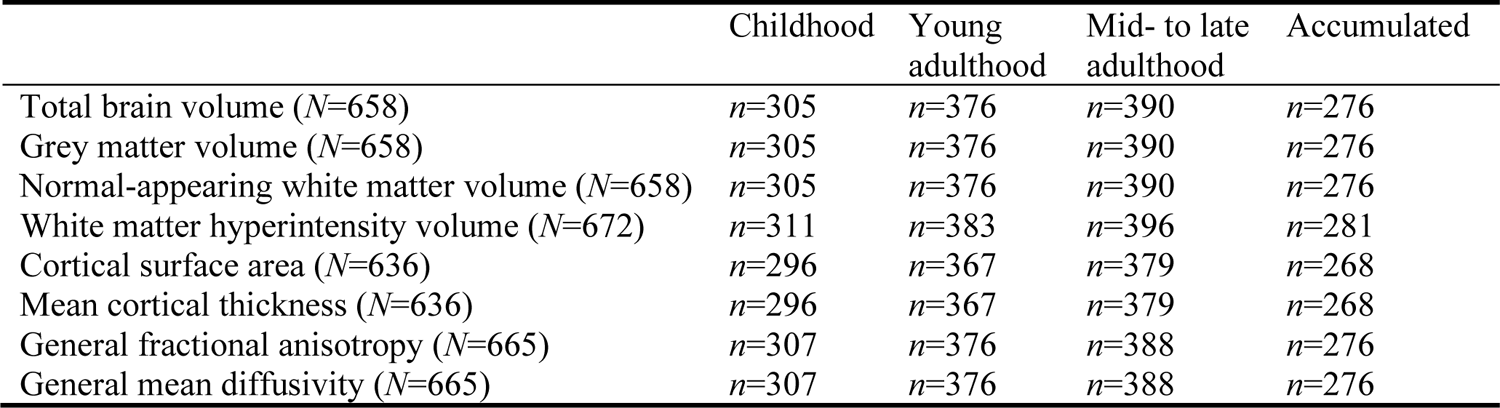
Number of complete cases for each neighbourhood deprivation exposure and global brain measure pairs.

|  | Childhood | Young adulthood | Mid- to late adulthood | Accumulated |
| --- | --- | --- | --- | --- |
| Total brain volume ($N=658$) | $n=305$ | $n=376$ | $n=390$ | $n=276$ |
| Grey matter volume ($N=658$) | $n=305$ | $n=376$ | $n=390$ | $n=276$ |
| Normal-appearing white matter volume ($N=658$) | $n=305$ | $n=376$ | $n=390$ | $n=276$ |
| White matter hyperintensity volume ($N=672$) | $n=311$ | $n=383$ | $n=396$ | $n=281$ |
| Cortical surface area ($N=636$) | $n=296$ | $n=367$ | $n=379$ | $n=268$ |
| Mean cortical thickness ($N=636$) | $n=296$ | $n=367$ | $n=379$ | $n=268$ |
| General fractional anisotropy ($N=665$) | $n=307$ | $n=376$ | $n=388$ | $n=276$ |
| General mean diffusivity ($N=665$) | $n=307$ | $n=376$ | $n=388$ | $n=276$ |

**Supplementary Table 4:**
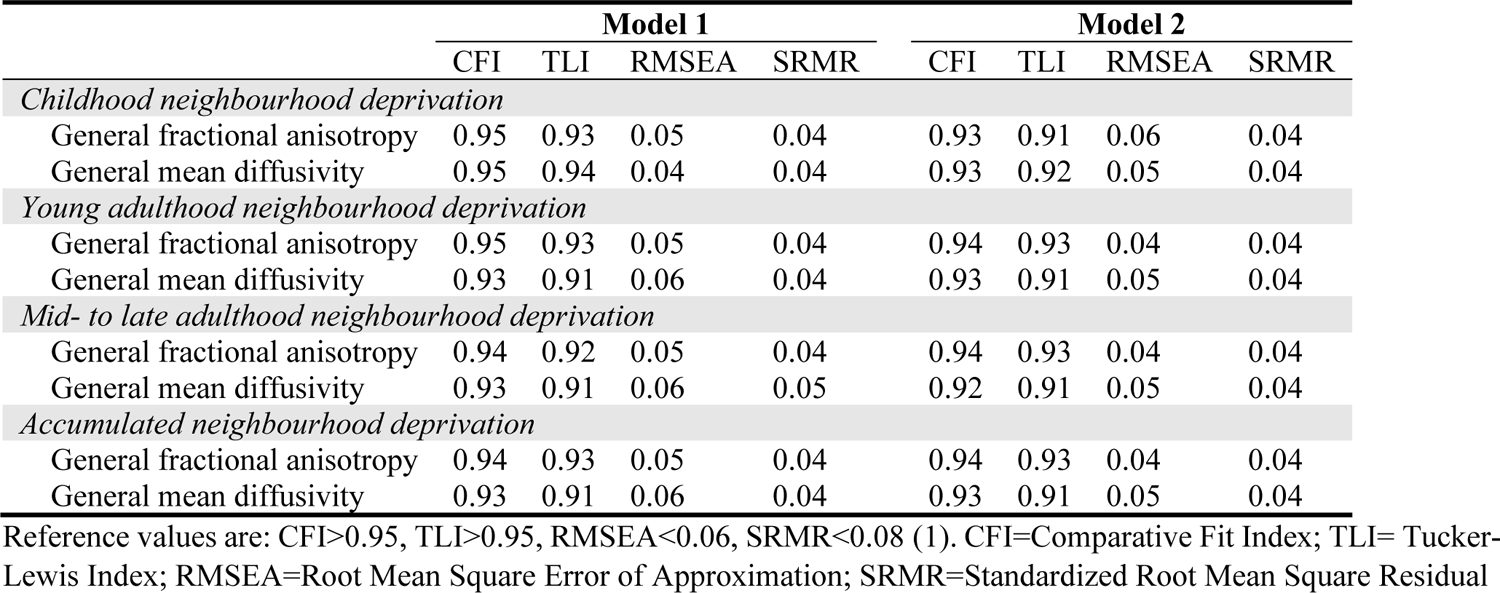
Model fit indices for general fractional anisotropy and general mean diffusivity in the main models

|  | **Model 1** |  |  |  | **Model 2** |  |  |  |
| --- | --- | --- | --- | --- | --- | --- | --- | --- |
|  | CFI | TLI | RMSEA | SRMR | CFI | TLI | RMSEA | SRMR |
| *Childhood neighbourhood deprivation* |  |  |  |  |  |  |  |  |
| General fractional anisotropy | 0.95 | 0.93 | 0.05 | 0.04 | 0.93 | 0.91 | 0.06 | 0.04 |
| General mean diffusivity | 0.95 | 0.94 | 0.04 | 0.04 | 0.93 | 0.92 | 0.05 | 0.04 |
| *Young adulthood neighbourhood deprivation* |  |  |  |  |  |  |  |  |
| General fractional anisotropy | 0.95 | 0.93 | 0.05 | 0.04 | 0.94 | 0.93 | 0.04 | 0.04 |
| General mean diffusivity | 0.93 | 0.91 | 0.06 | 0.04 | 0.93 | 0.91 | 0.05 | 0.04 |
| *Mid- to late adulthood neighbourhood deprivation* |  |  |  |  |  |  |  |  |
| General fractional anisotropy | 0.94 | 0.92 | 0.05 | 0.04 | 0.94 | 0.93 | 0.04 | 0.04 |
| General mean diffusivity | 0.93 | 0.91 | 0.06 | 0.05 | 0.92 | 0.91 | 0.05 | 0.04 |
| *Accumulated neighbourhood deprivation* |  |  |  |  |  |  |  |  |
| General fractional anisotropy | 0.94 | 0.93 | 0.05 | 0.04 | 0.94 | 0.93 | 0.04 | 0.04 |
| General mean diffusivity | 0.93 | 0.91 | 0.06 | 0.04 | 0.93 | 0.91 | 0.05 | 0.04 |

Reference values are: CFI>0.95, TLI>0.95, RMSEA<0.06, SRMR<0.08 (1). CFI=Comparative Fit Index; TLI= Tucker-Lewis Index; RMSEA=Root Mean Square Error of Approximation; SRMR=Standardized Root Mean Square Residual

**Supplementary Figure 1:**
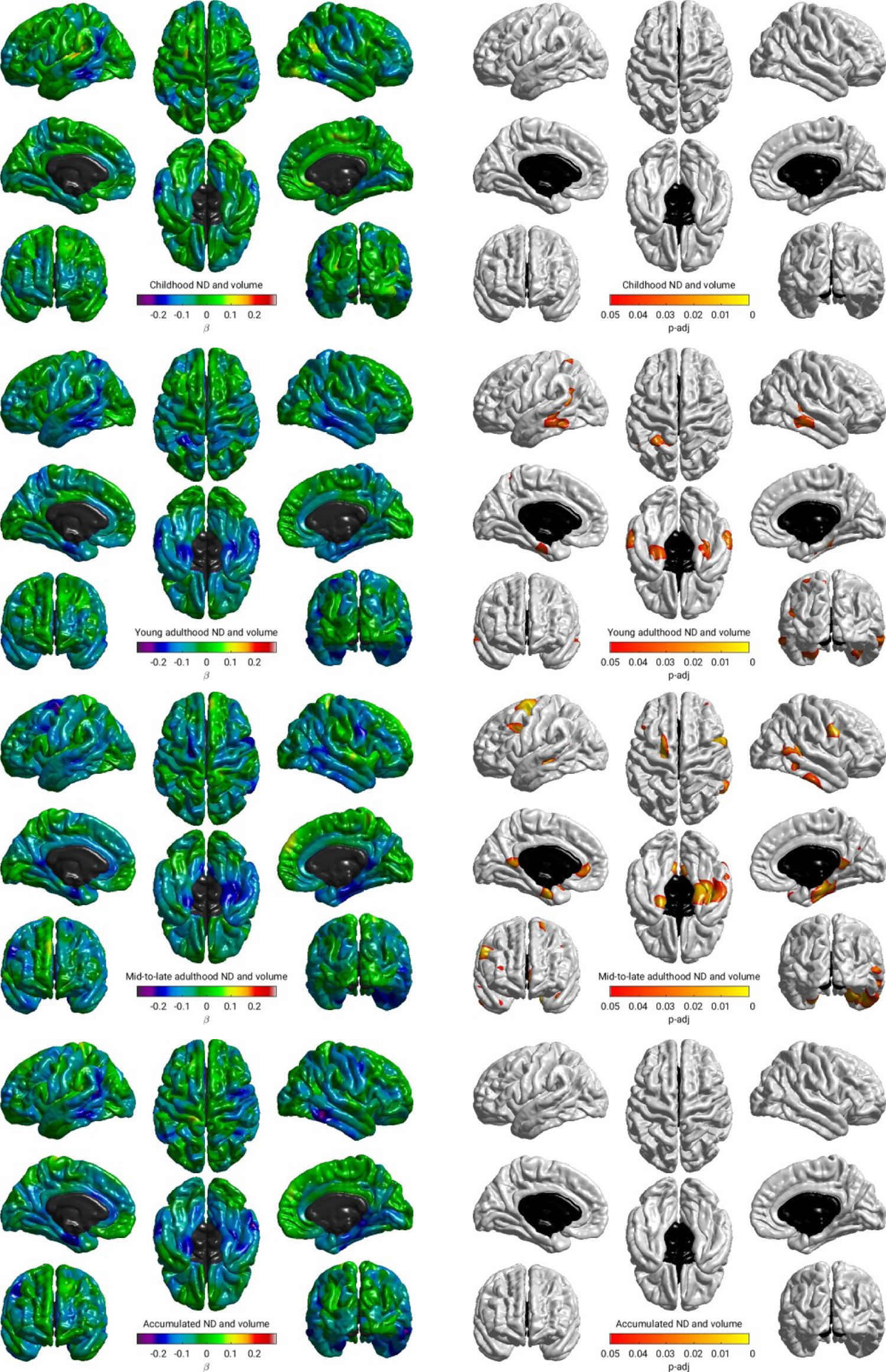
Regional associations between life-course models of neighbourhood deprivation and cortical volume in Model 1. Standardized coefficients were obtained in linear regression models fitted within the structural equation modelling framework applying full information maximum likelihood estimation. Sample size was *N*=622; pairwise complete observations were *n*=289, *n*=358, *n*=371 and *n*=262 for childhood, young adulthood, mid-to-late adulthood, and accumulated neighbourhood deprivation (ND), respectively. Models were adjusted for sex, age, and intracranial volume. Heatmaps show (from left to right): standardised betas and false discovery rate adjusted *p*-values (*pFDR* < 0.05). The non-cortical mask is shown in black.

**Supplementary Figure 2:**
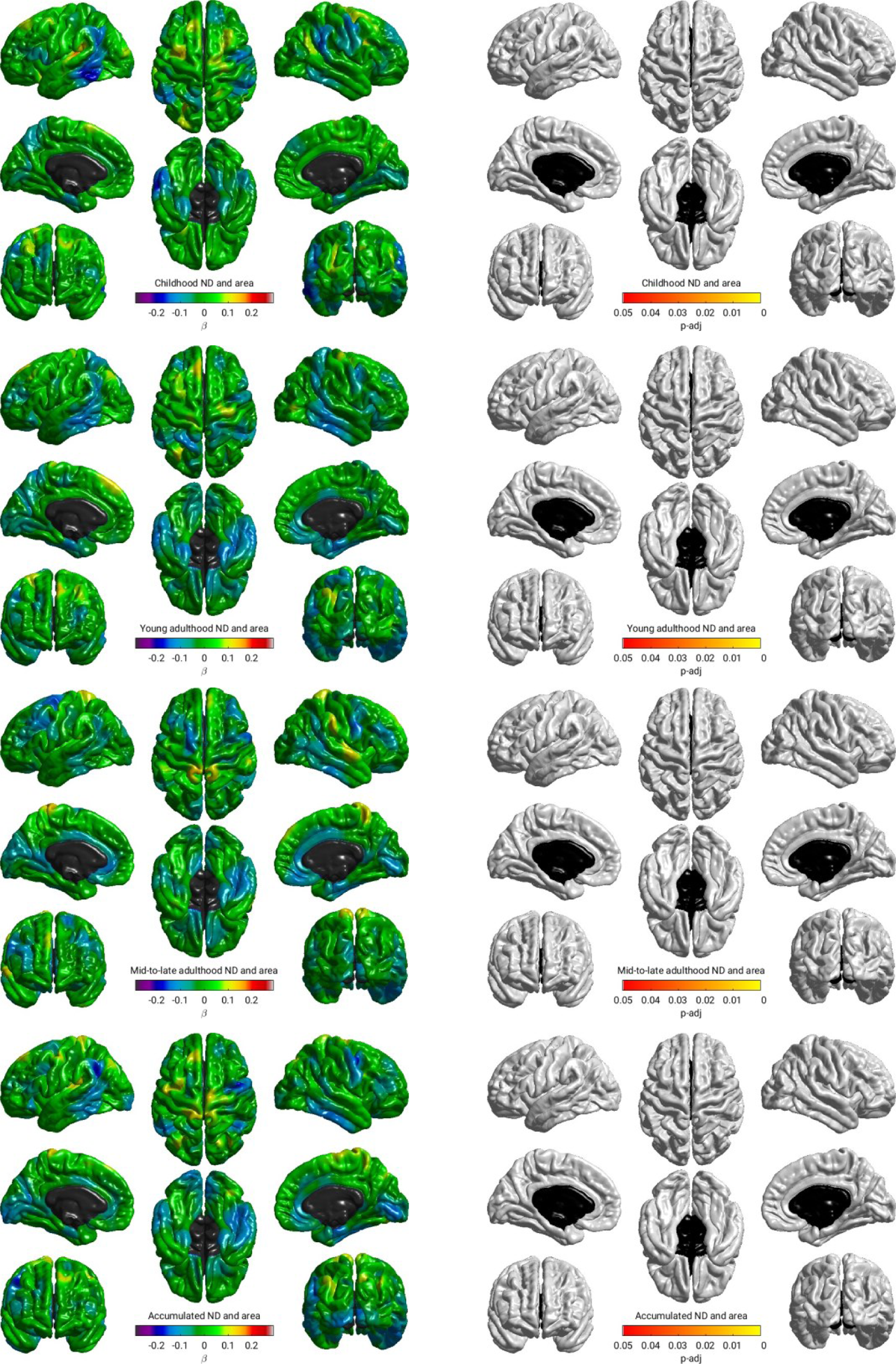
Regional associations between life-course models of neighbourhood deprivation and cortical surface area in Model 1. Standardized coefficients were obtained in linear regression models fitted within the structural equation modelling framework applying full information maximum likelihood estimation. Sample size was *N*=622; pairwise complete observations were *n*=289, *n*=358, *n*=371 and *n*=262 for childhood, young adulthood, mid-to-late adulthood, and accumulated neighbourhood deprivation (ND), respectively. Models were adjusted for sex, age, and intracranial volume. Heatmaps show (from left to right): standardised betas and false discovery rate adjusted *p*-values (*pFDR* < 0.05). The non-cortical mask is shown in black.

**Supplementary Figure 3:**
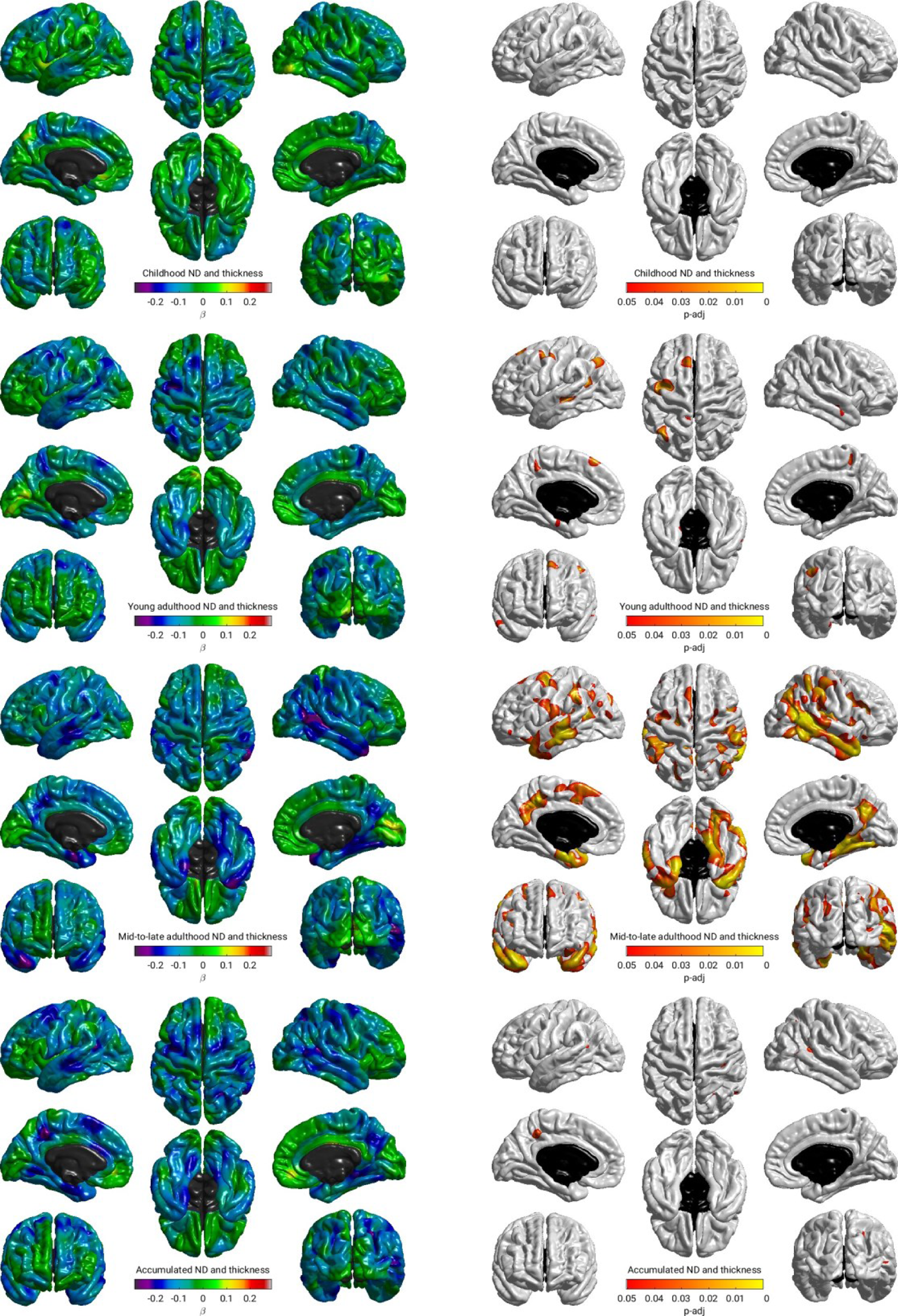
Regional associations between life-course models of neighbourhood deprivation and cortical thickness in Model 1. Standardized coefficients were obtained in linear regression models fitted within the structural equation modelling framework applying full information maximum likelihood estimation. Sample size was *N*=622; pairwise complete observations were *n*=289, *n*=358, *n*=371 and *n*=262 for childhood, young adulthood, mid-to-late adulthood, and accumulated neighbourhood deprivation (ND), respectively. Models were adjusted for sex, age, and intracranial volume. Heatmaps show (from left to right): standardised betas and false discovery rate adjusted *p*-values (*pFDR* < 0.05). The non-cortical mask is shown in black.

**Supplementary Figure 4:**
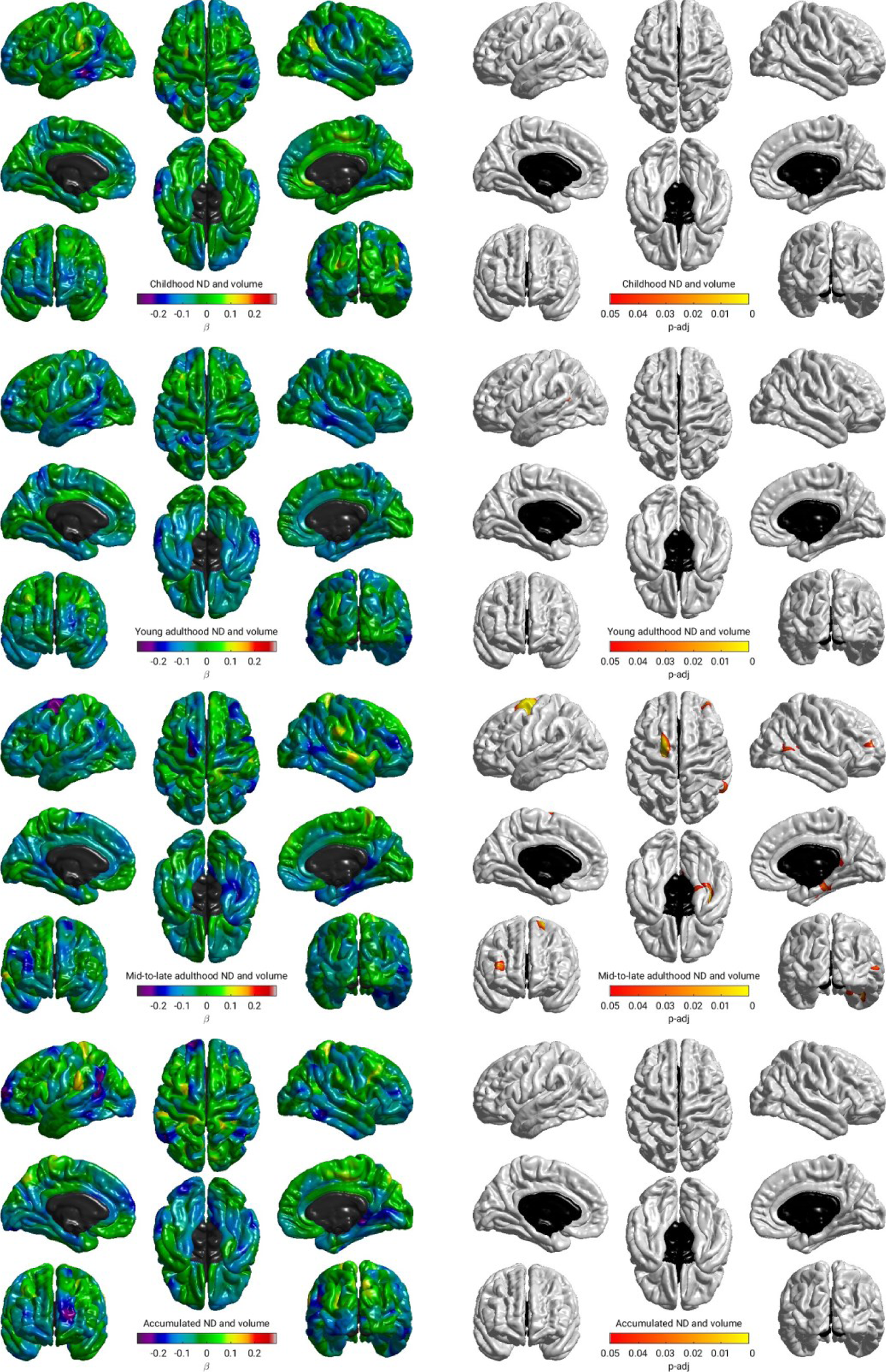
Regional associations between life-course models of neighbourhood deprivation and cortical volume in Model 2. Standardized coefficients were obtained in linear regression models fitted within the structural equation modelling framework applying full information maximum likelihood estimation. Sample size was *N*=622; pairwise complete observations were *n*=289, *n*=358, *n*=371 and *n*=262 for childhood, young adulthood, mid-to-late adulthood, and accumulated neighbourhood deprivation (ND), respectively. Models were adjusted for sex, age, intracranial volume, father’s occupational social class, APOE ε4 allele status. In addition, young adulthood models were adjusted for childhood IQ and years spent in education, and mid- to late adulthood/ accumulation models also for adult occupational social class. Heatmaps show (from left to right): standardised betas and false discovery rate adjusted *p*-values (*pFDR* < 0.05). The non-cortical mask is shown in black.

**Supplementary Figure 5:**
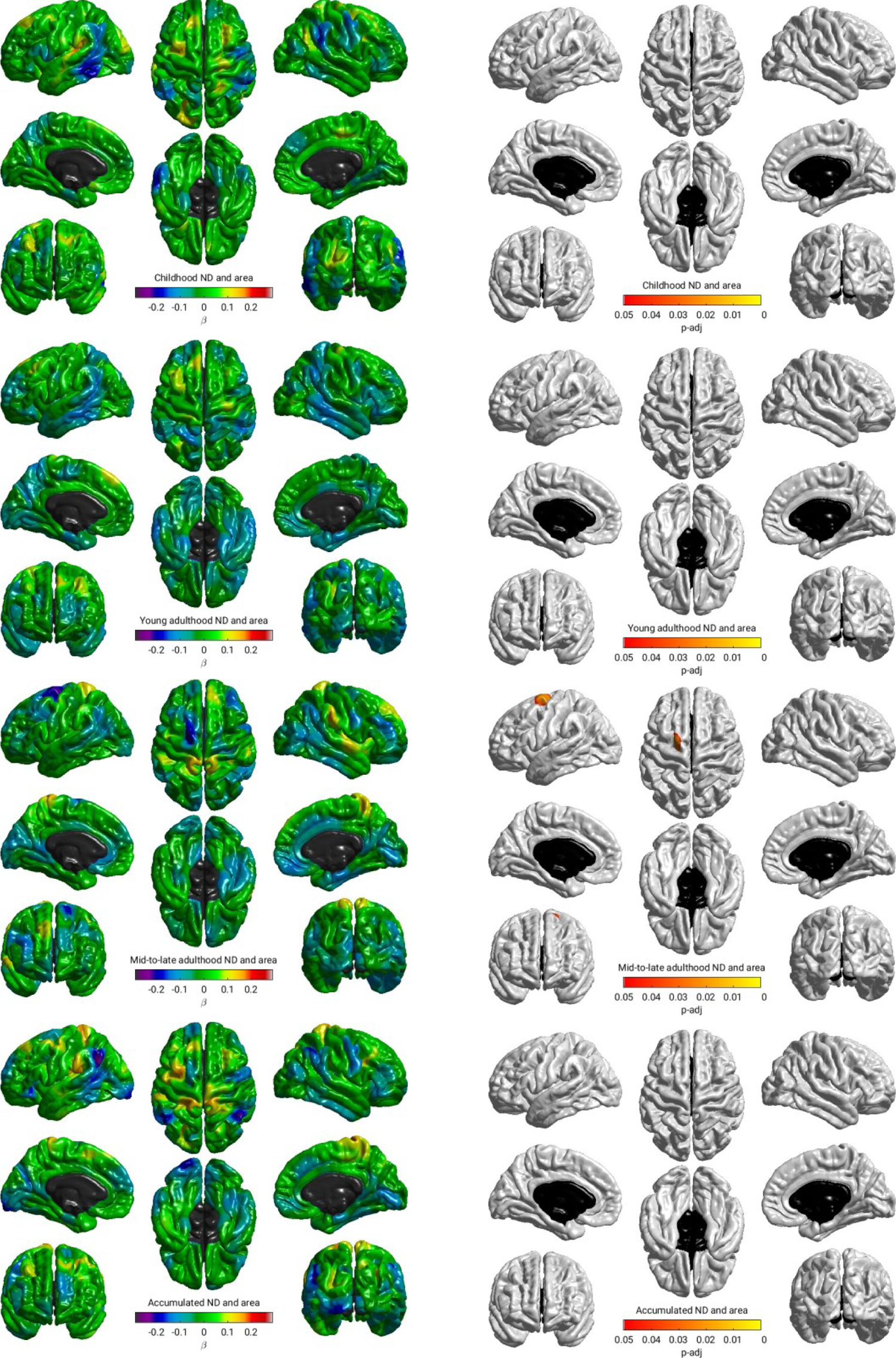
Regional associations between life-course models of neighbourhood deprivation and cortical surface area in Model 2. Standardized coefficients were obtained in linear regression models fitted within the structural equation modelling framework applying full information maximum likelihood estimation. Sample size was *N*=622; pairwise complete observations were *n*=289, *n*=358, *n*=371 and *n*=262 for childhood, young adulthood, mid-to-late adulthood, and accumulated neighbourhood deprivation (ND), respectively. Models were adjusted for sex, age, intracranial volume, father’s occupational social class, APOE ε4 allele status. In addition, young adulthood models were adjusted for childhood IQ and years spent in education, and mid- to late adulthood/ accumulation models also for adult occupational social class. Heatmaps show (from left to right): standardised betas and false discovery rate adjusted *p*-values (*pFDR* < 0.05). The non-cortical mask is shown in black.

**Supplementary Figure 6:**
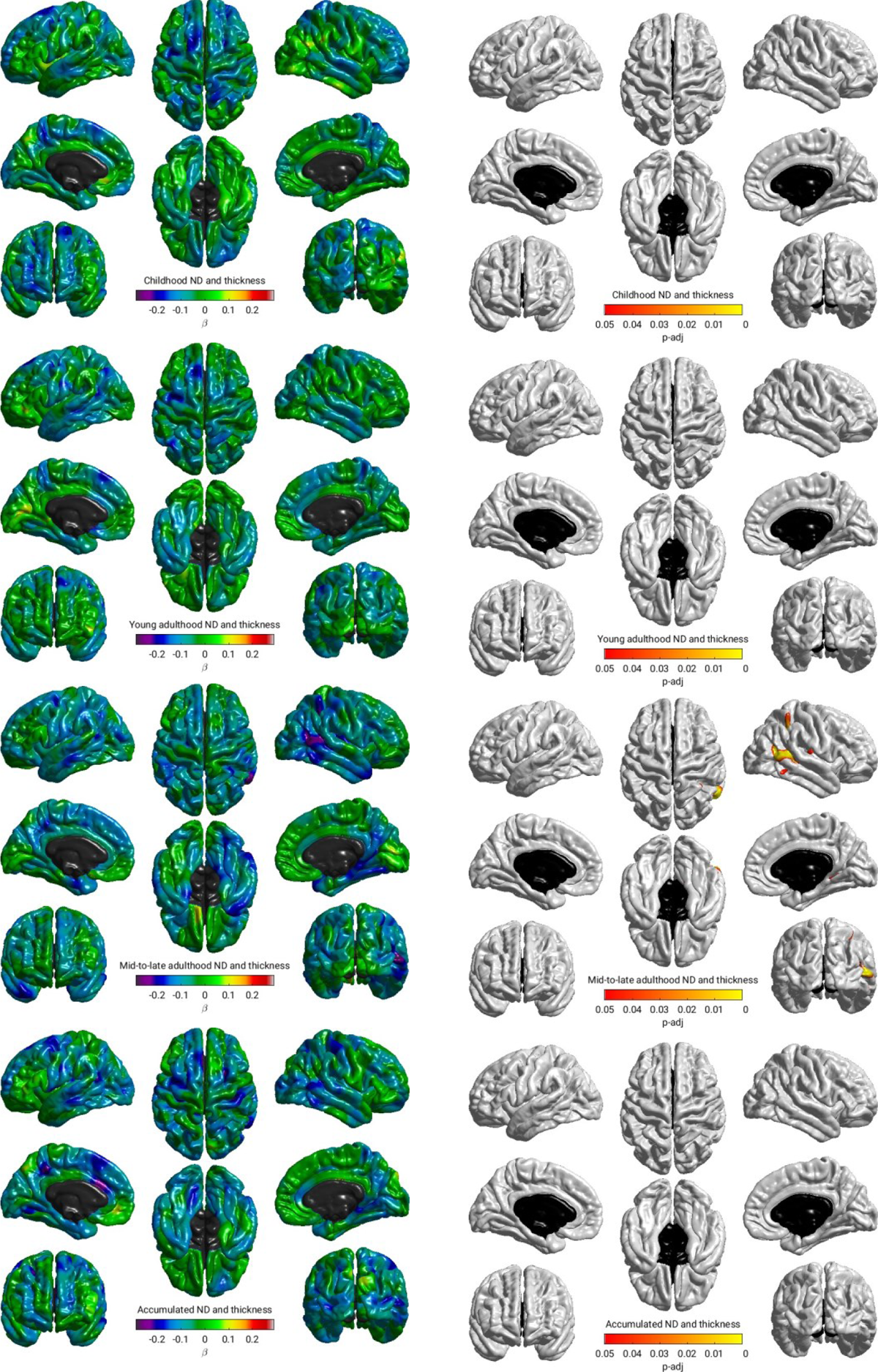
Regional associations between life-course models of neighbourhood deprivation and cortical thickness in Model 2. Standardized coefficients were obtained in linear regression models fitted within the structural equation modelling framework applying full information maximum likelihood estimation. Sample size was *N*=622; pairwise complete observations were *n*=289, *n*=358, *n*=371 and *n*=262 for childhood, young adulthood, mid-to-late adulthood, and accumulated neighbourhood deprivation (ND), respectively. Models were adjusted for sex, age, intracranial volume, father’s occupational social class, APOE ε4 allele status. In addition, young adulthood models were adjusted for childhood IQ and years spent in education, and mid- to late adulthood/ accumulation models also for adult occupational social class. Heatmaps show (from left to right): standardised betas and false discovery rate adjusted *p*-values (*pFDR* < 0.05). The non-cortical mask is shown in black.

**Supplementary Figure 7:**
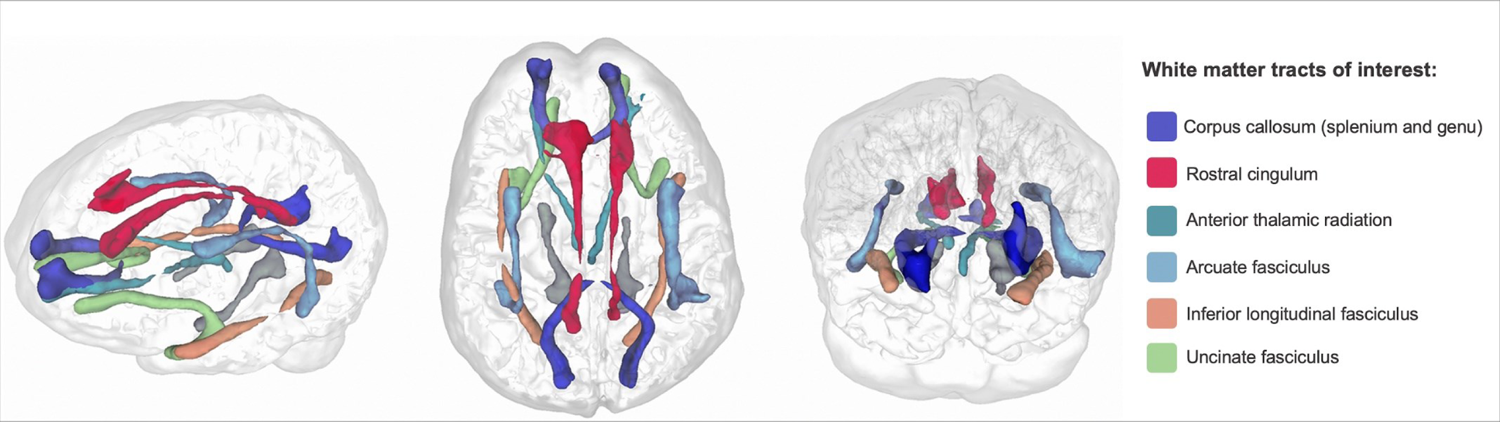
Glass brain plot locating white matter tracts used in the analyses. The ventral cingulum, coloured in grey, was not included in analyses as the rostral and ventral cingula are subdivisions of the same tract (an approach adopted previously in this cohort, e.g., (2))

**Supplementary Table 5:**
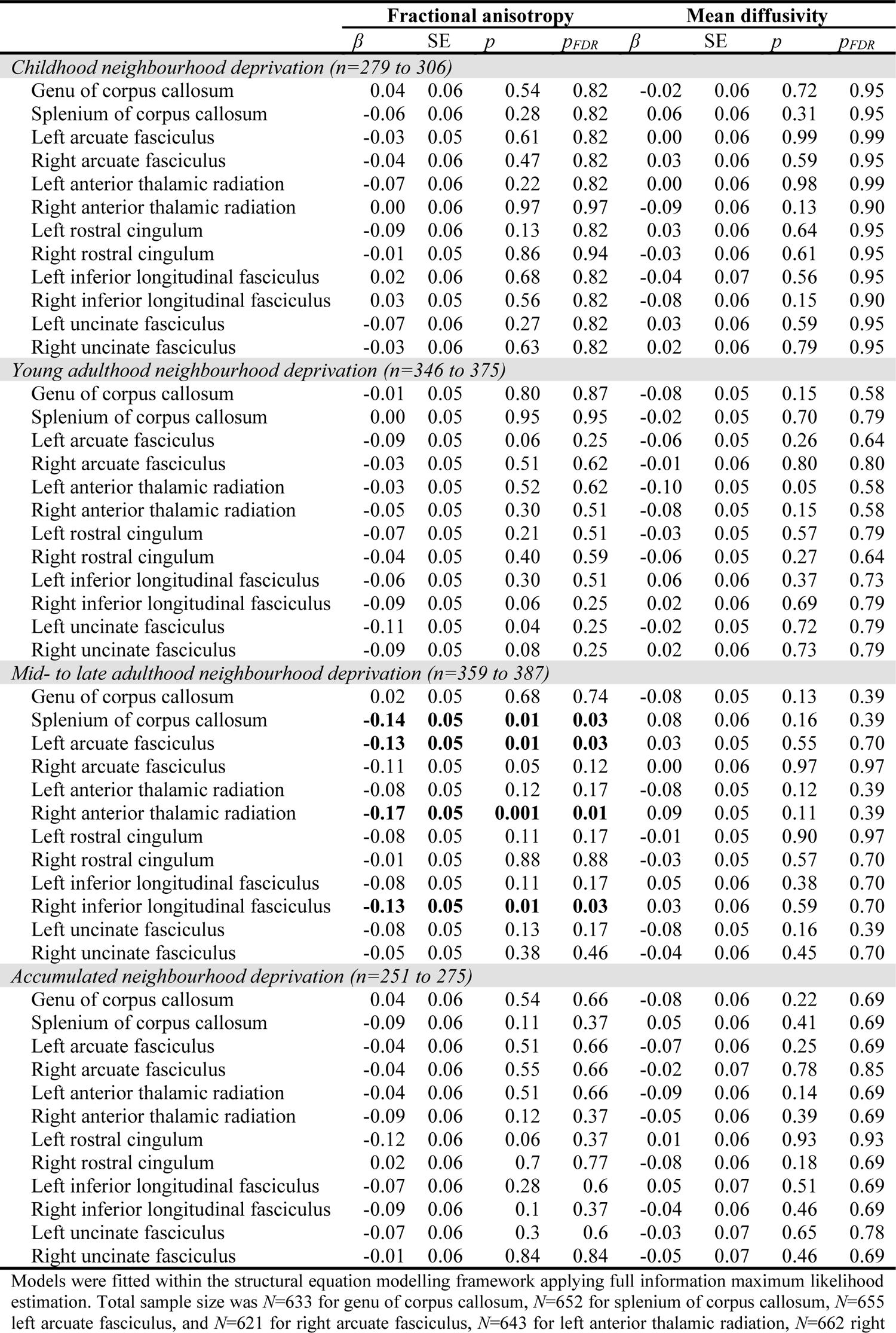
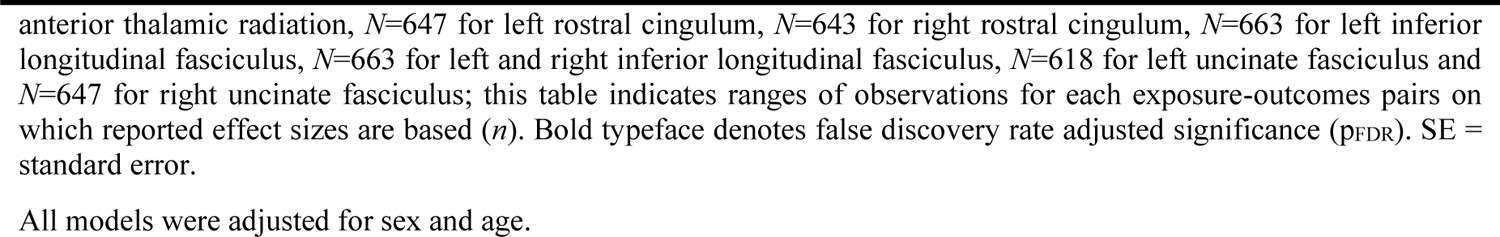
Association between life-course models of neighbourhood deprivation, fractional anisotropy and mean diffusivity in 12 white matter tracts in Model 1

|  | Fractional anisotropy |  |  |  | Mean diffusivity |  |  |  |
| --- | --- | --- | --- | --- | --- | --- | --- | --- |
| | $\beta$ | SE | $p$ | $p_{FDR}$ | $\beta$ | SE | $p$ | $p_{FDR}$ |
| *Childhood neighbourhood deprivation (n=279 to 306)* |  |  |  |  |  |  |  |  |
| Genu of corpus callosum | 0.04 | 0.06 | 0.54 | 0.82 | -0.02 | 0.06 | 0.72 | 0.95 |
| Splenium of corpus callosum | -0.06 | 0.06 | 0.28 | 0.82 | 0.06 | 0.06 | 0.31 | 0.95 |
| Left arcuate fasciculus | -0.03 | 0.05 | 0.61 | 0.82 | 0.00 | 0.06 | 0.99 | 0.99 |
| Right arcuate fasciculus | -0.04 | 0.06 | 0.47 | 0.82 | 0.03 | 0.06 | 0.59 | 0.95 |
| Left anterior thalamic radiation | -0.07 | 0.06 | 0.22 | 0.82 | 0.00 | 0.06 | 0.98 | 0.99 |
| Right anterior thalamic radiation | 0.00 | 0.06 | 0.97 | 0.97 | -0.09 | 0.06 | 0.13 | 0.90 |
| Left rostral cingulum | -0.09 | 0.06 | 0.13 | 0.82 | 0.03 | 0.06 | 0.64 | 0.95 |
| Right rostral cingulum | -0.01 | 0.05 | 0.86 | 0.94 | -0.03 | 0.06 | 0.61 | 0.95 |
| Left inferior longitudinal fasciculus | 0.02 | 0.06 | 0.68 | 0.82 | -0.04 | 0.07 | 0.56 | 0.95 |
| Right inferior longitudinal fasciculus | 0.03 | 0.05 | 0.56 | 0.82 | -0.08 | 0.06 | 0.15 | 0.90 |
| Left uncinate fasciculus | -0.07 | 0.06 | 0.27 | 0.82 | 0.03 | 0.06 | 0.59 | 0.95 |
| Right uncinate fasciculus | -0.03 | 0.06 | 0.63 | 0.82 | 0.02 | 0.06 | 0.79 | 0.95 |
| *Young adulthood neighbourhood deprivation (n=346 to 375)* |  |  |  |  |  |  |  |  |
| Genu of corpus callosum | -0.01 | 0.05 | 0.80 | 0.87 | -0.08 | 0.05 | 0.15 | 0.58 |
| Splenium of corpus callosum | 0.00 | 0.05 | 0.95 | 0.95 | -0.02 | 0.05 | 0.70 | 0.79 |
| Left arcuate fasciculus | -0.09 | 0.05 | 0.06 | 0.25 | -0.06 | 0.05 | 0.26 | 0.64 |
| Right arcuate fasciculus | -0.03 | 0.05 | 0.51 | 0.62 | -0.01 | 0.06 | 0.80 | 0.80 |
| Left anterior thalamic radiation | -0.03 | 0.05 | 0.52 | 0.62 | -0.10 | 0.05 | 0.05 | 0.58 |
| Right anterior thalamic radiation | -0.05 | 0.05 | 0.30 | 0.51 | -0.08 | 0.05 | 0.15 | 0.58 |
| Left rostral cingulum | -0.07 | 0.05 | 0.21 | 0.51 | -0.03 | 0.05 | 0.57 | 0.79 |
| Right rostral cingulum | -0.04 | 0.05 | 0.40 | 0.59 | -0.06 | 0.05 | 0.27 | 0.64 |
| Left inferior longitudinal fasciculus | -0.06 | 0.05 | 0.30 | 0.51 | 0.06 | 0.06 | 0.37 | 0.73 |
| Right inferior longitudinal fasciculus | -0.09 | 0.05 | 0.06 | 0.25 | 0.02 | 0.06 | 0.69 | 0.79 |
| Left uncinate fasciculus | -0.11 | 0.05 | 0.04 | 0.25 | -0.02 | 0.05 | 0.72 | 0.79 |
| Right uncinate fasciculus | -0.09 | 0.05 | 0.08 | 0.25 | 0.02 | 0.06 | 0.73 | 0.79 |
| *Mid- to late adulthood neighbourhood deprivation (n=359 to 387)* |  |  |  |  |  |  |  |  |
| Genu of corpus callosum | 0.02 | 0.05 | 0.68 | 0.74 | -0.08 | 0.05 | 0.13 | 0.39 |
| Splenium of corpus callosum | **-0.14** | **0.05** | **0.01** | **0.03** | 0.08 | 0.06 | 0.16 | 0.39 |
| Left arcuate fasciculus | **-0.13** | **0.05** | **0.01** | **0.03** | 0.03 | 0.05 | 0.55 | 0.70 |
| Right arcuate fasciculus | -0.11 | 0.05 | 0.05 | 0.12 | 0.00 | 0.06 | 0.97 | 0.97 |
| Left anterior thalamic radiation | -0.08 | 0.05 | 0.12 | 0.17 | -0.08 | 0.05 | 0.12 | 0.39 |
| Right anterior thalamic radiation | **-0.17** | **0.05** | **0.001** | **0.01** | 0.09 | 0.05 | 0.11 | 0.39 |
| Left rostral cingulum | -0.08 | 0.05 | 0.11 | 0.17 | -0.01 | 0.05 | 0.90 | 0.97 |
| Right rostral cingulum | -0.01 | 0.05 | 0.88 | 0.88 | -0.03 | 0.05 | 0.57 | 0.70 |
| Left inferior longitudinal fasciculus | -0.08 | 0.05 | 0.11 | 0.17 | 0.05 | 0.06 | 0.38 | 0.70 |
| Right inferior longitudinal fasciculus | **-0.13** | **0.05** | **0.01** | **0.03** | 0.03 | 0.06 | 0.59 | 0.70 |
| Left uncinate fasciculus | -0.08 | 0.05 | 0.13 | 0.17 | -0.08 | 0.05 | 0.16 | 0.39 |
| Right uncinate fasciculus | -0.05 | 0.05 | 0.38 | 0.46 | -0.04 | 0.06 | 0.45 | 0.70 |
| *Accumulated neighbourhood deprivation (n=251 to 275)* |  |  |  |  |  |  |  |  |
| Genu of corpus callosum | 0.04 | 0.06 | 0.54 | 0.66 | -0.08 | 0.06 | 0.22 | 0.69 |
| Splenium of corpus callosum | -0.09 | 0.06 | 0.11 | 0.37 | 0.05 | 0.06 | 0.41 | 0.69 |
| Left arcuate fasciculus | -0.04 | 0.06 | 0.51 | 0.66 | -0.07 | 0.06 | 0.25 | 0.69 |
| Right arcuate fasciculus | -0.04 | 0.06 | 0.55 | 0.66 | -0.02 | 0.07 | 0.78 | 0.85 |
| Left anterior thalamic radiation | -0.04 | 0.06 | 0.51 | 0.66 | -0.09 | 0.06 | 0.14 | 0.69 |
| Right anterior thalamic radiation | -0.09 | 0.06 | 0.12 | 0.37 | -0.05 | 0.06 | 0.39 | 0.69 |
| Left rostral cingulum | -0.12 | 0.06 | 0.06 | 0.37 | 0.01 | 0.06 | 0.93 | 0.93 |
| Right rostral cingulum | 0.02 | 0.06 | 0.7 | 0.77 | -0.08 | 0.06 | 0.18 | 0.69 |
| Left inferior longitudinal fasciculus | -0.07 | 0.06 | 0.28 | 0.6 | 0.05 | 0.07 | 0.51 | 0.69 |
| Right inferior longitudinal fasciculus | -0.09 | 0.06 | 0.1 | 0.37 | -0.04 | 0.06 | 0.46 | 0.69 |
| Left uncinate fasciculus | -0.07 | 0.06 | 0.3 | 0.6 | -0.03 | 0.07 | 0.65 | 0.78 |
| Right uncinate fasciculus | -0.01 | 0.06 | 0.84 | 0.84 | -0.05 | 0.07 | 0.46 | 0.69 |

Models were fitted within the structural equation modelling framework applying full information maximum likelihood estimation. Total sample size was $N=633$ for genu of corpus callosum, $N=652$ for splenium of corpus callosum, $N=655$ left arcuate fasciculus, and $N=621$ for right arcuate fasciculus, $N=643$ for left anterior thalamic radiation, $N=662$ right
anterior thalamic radiation, $N=647$ for left rostral cingulum, $N=643$ for right rostral cingulum, $N=663$ for left inferior longitudinal fasciculus, $N=663$ for left and right inferior longitudinal fasciculus, $N=618$ for left uncinate fasciculus and $N=647$ for right uncinate fasciculus; this table indicates ranges of observations for each exposure-outcomes pairs on which reported effect sizes are based ( $n$ ). Bold typeface denotes false discovery rate adjusted significance ( $p_{FDR}$ ). SE = standard error.
All models were adjusted for sex and age.

**Supplementary Table 6:**
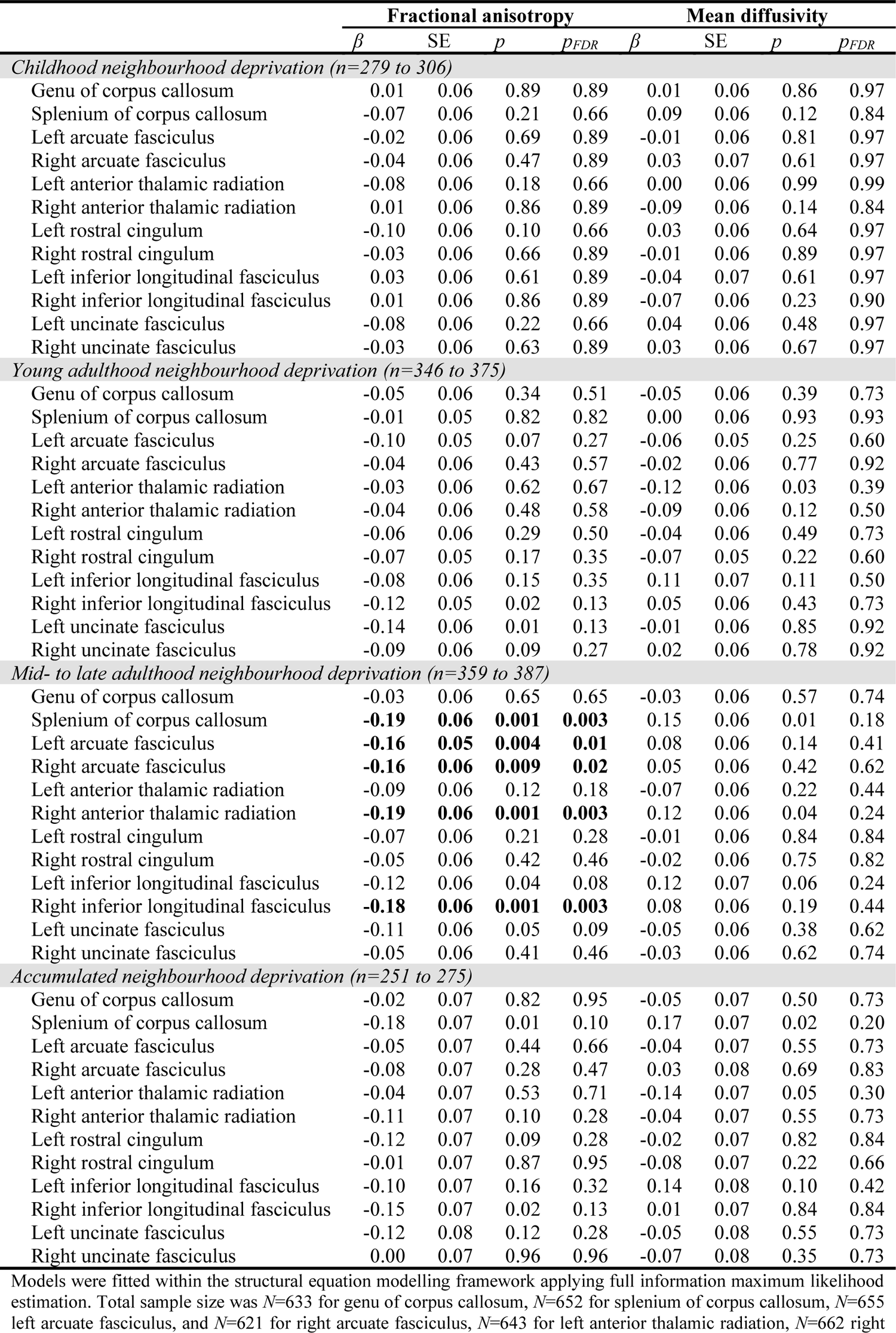
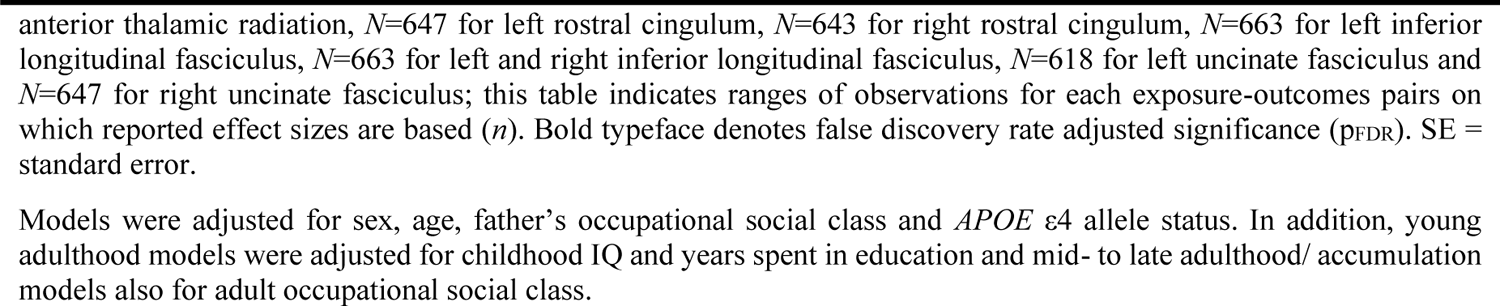
Association between life-course models of neighbourhood deprivation, fractional anisotropy and mean diffusivity in 12 white matter tracts in Model 2

|  | Fractional anisotropy |  |  |  | Mean diffusivity |  |  |  |
| --- | --- | --- | --- | --- | --- | --- | --- | --- |
| | $\beta$ | SE | $p$ | $p_{FDR}$ | $\beta$ | SE | $p$ | $p_{FDR}$ |
| *Childhood neighbourhood deprivation (n=279 to 306)* |  |  |  |  |  |  |  |  |
| Genu of corpus callosum | 0.01 | 0.06 | 0.89 | 0.89 | 0.01 | 0.06 | 0.86 | 0.97 |
| Splenium of corpus callosum | -0.07 | 0.06 | 0.21 | 0.66 | 0.09 | 0.06 | 0.12 | 0.84 |
| Left arcuate fasciculus | -0.02 | 0.06 | 0.69 | 0.89 | -0.01 | 0.06 | 0.81 | 0.97 |
| Right arcuate fasciculus | -0.04 | 0.06 | 0.47 | 0.89 | 0.03 | 0.07 | 0.61 | 0.97 |
| Left anterior thalamic radiation | -0.08 | 0.06 | 0.18 | 0.66 | 0.00 | 0.06 | 0.99 | 0.99 |
| Right anterior thalamic radiation | 0.01 | 0.06 | 0.86 | 0.89 | -0.09 | 0.06 | 0.14 | 0.84 |
| Left rostral cingulum | -0.10 | 0.06 | 0.10 | 0.66 | 0.03 | 0.06 | 0.64 | 0.97 |
| Right rostral cingulum | -0.03 | 0.06 | 0.66 | 0.89 | -0.01 | 0.06 | 0.89 | 0.97 |
| Left inferior longitudinal fasciculus | 0.03 | 0.06 | 0.61 | 0.89 | -0.04 | 0.07 | 0.61 | 0.97 |
| Right inferior longitudinal fasciculus | 0.01 | 0.06 | 0.86 | 0.89 | -0.07 | 0.06 | 0.23 | 0.90 |
| Left uncinate fasciculus | -0.08 | 0.06 | 0.22 | 0.66 | 0.04 | 0.06 | 0.48 | 0.97 |
| Right uncinate fasciculus | -0.03 | 0.06 | 0.63 | 0.89 | 0.03 | 0.06 | 0.67 | 0.97 |
| *Young adulthood neighbourhood deprivation (n=346 to 375)* |  |  |  |  |  |  |  |  |
| Genu of corpus callosum | -0.05 | 0.06 | 0.34 | 0.51 | -0.05 | 0.06 | 0.39 | 0.73 |
| Splenium of corpus callosum | -0.01 | 0.05 | 0.82 | 0.82 | 0.00 | 0.06 | 0.93 | 0.93 |
| Left arcuate fasciculus | -0.10 | 0.05 | 0.07 | 0.27 | -0.06 | 0.05 | 0.25 | 0.60 |
| Right arcuate fasciculus | -0.04 | 0.06 | 0.43 | 0.57 | -0.02 | 0.06 | 0.77 | 0.92 |
| Left anterior thalamic radiation | -0.03 | 0.06 | 0.62 | 0.67 | -0.12 | 0.06 | 0.03 | 0.39 |
| Right anterior thalamic radiation | -0.04 | 0.06 | 0.48 | 0.58 | -0.09 | 0.06 | 0.12 | 0.50 |
| Left rostral cingulum | -0.06 | 0.06 | 0.29 | 0.50 | -0.04 | 0.06 | 0.49 | 0.73 |
| Right rostral cingulum | -0.07 | 0.05 | 0.17 | 0.35 | -0.07 | 0.05 | 0.22 | 0.60 |
| Left inferior longitudinal fasciculus | -0.08 | 0.06 | 0.15 | 0.35 | 0.11 | 0.07 | 0.11 | 0.50 |
| Right inferior longitudinal fasciculus | -0.12 | 0.05 | 0.02 | 0.13 | 0.05 | 0.06 | 0.43 | 0.73 |
| Left uncinate fasciculus | -0.14 | 0.06 | 0.01 | 0.13 | -0.01 | 0.06 | 0.85 | 0.92 |
| Right uncinate fasciculus | -0.09 | 0.06 | 0.09 | 0.27 | 0.02 | 0.06 | 0.78 | 0.92 |
| *Mid- to late adulthood neighbourhood deprivation (n=359 to 387)* |  |  |  |  |  |  |  |  |
| Genu of corpus callosum | -0.03 | 0.06 | 0.65 | 0.65 | -0.03 | 0.06 | 0.57 | 0.74 |
| Splenium of corpus callosum | **-0.19** | **0.06** | **0.001** | **0.003** | 0.15 | 0.06 | 0.01 | 0.18 |
| Left arcuate fasciculus | **-0.16** | **0.05** | **0.004** | **0.01** | 0.08 | 0.06 | 0.14 | 0.41 |
| Right arcuate fasciculus | **-0.16** | **0.06** | **0.009** | **0.02** | 0.05 | 0.06 | 0.42 | 0.62 |
| Left anterior thalamic radiation | -0.09 | 0.06 | 0.12 | 0.18 | -0.07 | 0.06 | 0.22 | 0.44 |
| Right anterior thalamic radiation | **-0.19** | **0.06** | **0.001** | **0.003** | 0.12 | 0.06 | 0.04 | 0.24 |
| Left rostral cingulum | -0.07 | 0.06 | 0.21 | 0.28 | -0.01 | 0.06 | 0.84 | 0.84 |
| Right rostral cingulum | -0.05 | 0.06 | 0.42 | 0.46 | -0.02 | 0.06 | 0.75 | 0.82 |
| Left inferior longitudinal fasciculus | -0.12 | 0.06 | 0.04 | 0.08 | 0.12 | 0.07 | 0.06 | 0.24 |
| Right inferior longitudinal fasciculus | **-0.18** | **0.06** | **0.001** | **0.003** | 0.08 | 0.06 | 0.19 | 0.44 |
| Left uncinate fasciculus | -0.11 | 0.06 | 0.05 | 0.09 | -0.05 | 0.06 | 0.38 | 0.62 |
| Right uncinate fasciculus | -0.05 | 0.06 | 0.41 | 0.46 | -0.03 | 0.06 | 0.62 | 0.74 |
| *Accumulated neighbourhood deprivation (n=251 to 275)* |  |  |  |  |  |  |  |  |
| Genu of corpus callosum | -0.02 | 0.07 | 0.82 | 0.95 | -0.05 | 0.07 | 0.50 | 0.73 |
| Splenium of corpus callosum | -0.18 | 0.07 | 0.01 | 0.10 | 0.17 | 0.07 | 0.02 | 0.20 |
| Left arcuate fasciculus | -0.05 | 0.07 | 0.44 | 0.66 | -0.04 | 0.07 | 0.55 | 0.73 |
| Right arcuate fasciculus | -0.08 | 0.07 | 0.28 | 0.47 | 0.03 | 0.08 | 0.69 | 0.83 |
| Left anterior thalamic radiation | -0.04 | 0.07 | 0.53 | 0.71 | -0.14 | 0.07 | 0.05 | 0.30 |
| Right anterior thalamic radiation | -0.11 | 0.07 | 0.10 | 0.28 | -0.04 | 0.07 | 0.55 | 0.73 |
| Left rostral cingulum | -0.12 | 0.07 | 0.09 | 0.28 | -0.02 | 0.07 | 0.82 | 0.84 |
| Right rostral cingulum | -0.01 | 0.07 | 0.87 | 0.95 | -0.08 | 0.07 | 0.22 | 0.66 |
| Left inferior longitudinal fasciculus | -0.10 | 0.07 | 0.16 | 0.32 | 0.14 | 0.08 | 0.10 | 0.42 |
| Right inferior longitudinal fasciculus | -0.15 | 0.07 | 0.02 | 0.13 | 0.01 | 0.07 | 0.84 | 0.84 |
| Left uncinate fasciculus | -0.12 | 0.08 | 0.12 | 0.28 | -0.05 | 0.08 | 0.55 | 0.73 |
| Right uncinate fasciculus | 0.00 | 0.07 | 0.96 | 0.96 | -0.07 | 0.08 | 0.35 | 0.73 |

Models were fitted within the structural equation modelling framework applying full information maximum likelihood estimation. Total sample size was $N=633$ for genu of corpus callosum, $N=652$ for splenium of corpus callosum, $N=655$ left arcuate fasciculus, and $N=621$ for right arcuate fasciculus, $N=643$ for left anterior thalamic radiation, $N=662$ right
anterior thalamic radiation, $N=647$ for left rostral cingulum, $N=643$ for right rostral cingulum, $N=663$ for left inferior longitudinal fasciculus, $N=663$ for left and right inferior longitudinal fasciculus, $N=618$ for left uncinate fasciculus and $N=647$ for right uncinate fasciculus; this table indicates ranges of observations for each exposure-outcomes pairs on which reported effect sizes are based ( $n$ ). Bold typeface denotes false discovery rate adjusted significance ( $p_{FDR}$ ). SE = standard error.
Models were adjusted for sex, age, father's occupational social class and *APOE* $\epsilon 4$ allele status. In addition, young adulthood models were adjusted for childhood IQ and years spent in education and mid- to late adulthood/ accumulation models also for adult occupational social class.

**Supplementary Table 7:**
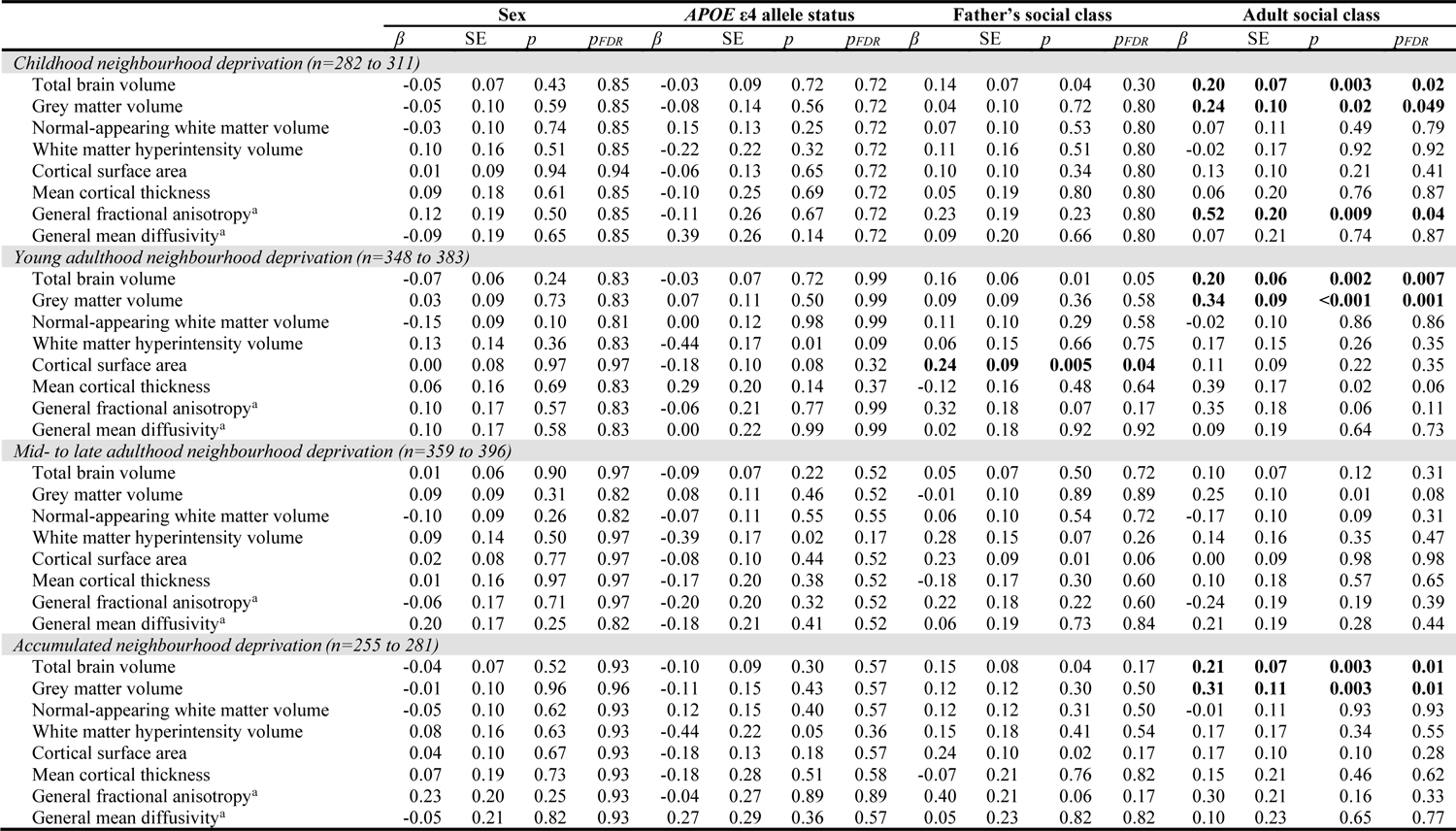
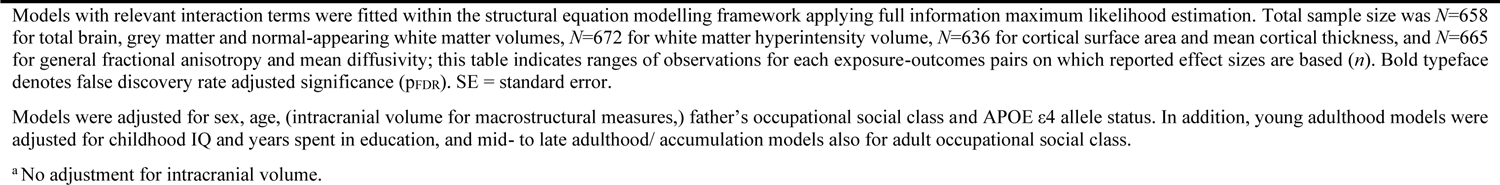
Interaction of neighbourhood deprivation with sex, *APOE* ε4 allele status and adult occupational social class and global brain outcomes

| | Sex | | | | *APOE* $\epsilon 4$ allele status | | | | Father's social class | | | | Adult social class | | | |
| --- | --- | --- | --- | --- | --- | --- | --- | --- | --- | --- | --- | --- | --- | --- | --- | --- |
| | $\beta$ | SE | *p* | *p*<sub>FDR</sub> | $\beta$ | SE | *p* | *p*<sub>FDR</sub> | $\beta$ | SE | *p* | *p*<sub>FDR</sub> | $\beta$ | SE | *p* | *p*<sub>FDR</sub> |
| *Childhood neighbourhood deprivation (n=282 to 311)* |  |  |  |  |  |  |  |  |  |  |  |  |  |  |  |  |
| Total brain volume | -0.05 | 0.07 | 0.43 | 0.85 | -0.03 | 0.09 | 0.72 | 0.72 | 0.14 | 0.07 | 0.04 | 0.30 | **0.20** | **0.07** | **0.003** | **0.02** |
| Grey matter volume | -0.05 | 0.10 | 0.59 | 0.85 | -0.08 | 0.14 | 0.56 | 0.72 | 0.04 | 0.10 | 0.72 | 0.80 | **0.24** | **0.10** | **0.02** | **0.049** |
| Normal-appearing white matter volume | -0.03 | 0.10 | 0.74 | 0.85 | 0.15 | 0.13 | 0.25 | 0.72 | 0.07 | 0.10 | 0.53 | 0.80 | 0.07 | 0.11 | 0.49 | 0.79 |
| White matter hyperintensity volume | 0.10 | 0.16 | 0.51 | 0.85 | -0.22 | 0.22 | 0.32 | 0.72 | 0.11 | 0.16 | 0.51 | 0.80 | -0.02 | 0.17 | 0.92 | 0.92 |
| Cortical surface area | 0.01 | 0.09 | 0.94 | 0.94 | -0.06 | 0.13 | 0.65 | 0.72 | 0.10 | 0.10 | 0.34 | 0.80 | 0.13 | 0.10 | 0.21 | 0.41 |
| Mean cortical thickness | 0.09 | 0.18 | 0.61 | 0.85 | -0.10 | 0.25 | 0.69 | 0.72 | 0.05 | 0.19 | 0.80 | 0.80 | 0.06 | 0.20 | 0.76 | 0.87 |
| General fractional anisotropy<sup>a</sup> | 0.12 | 0.19 | 0.50 | 0.85 | -0.11 | 0.26 | 0.67 | 0.72 | 0.23 | 0.19 | 0.23 | 0.80 | **0.52** | **0.20** | **0.009** | **0.04** |
| General mean diffusivity<sup>a</sup> | -0.09 | 0.19 | 0.65 | 0.85 | 0.39 | 0.26 | 0.14 | 0.72 | 0.09 | 0.20 | 0.66 | 0.80 | 0.07 | 0.21 | 0.74 | 0.87 |
| *Young adulthood neighbourhood deprivation (n=348 to 383)* |  |  |  |  |  |  |  |  |  |  |  |  |  |  |  |  |
| Total brain volume | -0.07 | 0.06 | 0.24 | 0.83 | -0.03 | 0.07 | 0.72 | 0.99 | 0.16 | 0.06 | 0.01 | 0.05 | **0.20** | **0.06** | **0.002** | **0.007** |
| Grey matter volume | 0.03 | 0.09 | 0.73 | 0.83 | 0.07 | 0.11 | 0.50 | 0.99 | 0.09 | 0.09 | 0.36 | 0.58 | **0.34** | **0.09** | **<0.001** | **0.001** |
| Normal-appearing white matter volume | -0.15 | 0.09 | 0.10 | 0.81 | 0.00 | 0.12 | 0.98 | 0.99 | 0.11 | 0.10 | 0.29 | 0.58 | -0.02 | 0.10 | 0.86 | 0.86 |
| White matter hyperintensity volume | 0.13 | 0.14 | 0.36 | 0.83 | -0.44 | 0.17 | 0.01 | 0.09 | 0.06 | 0.15 | 0.66 | 0.75 | 0.17 | 0.15 | 0.26 | 0.35 |
| Cortical surface area | 0.00 | 0.08 | 0.97 | 0.97 | -0.18 | 0.10 | 0.08 | 0.32 | **0.24** | **0.09** | **0.005** | **0.04** | 0.11 | 0.09 | 0.22 | 0.35 |
| Mean cortical thickness | 0.06 | 0.16 | 0.69 | 0.83 | 0.29 | 0.20 | 0.14 | 0.37 | -0.12 | 0.16 | 0.48 | 0.64 | 0.39 | 0.17 | 0.02 | 0.06 |
| General fractional anisotropy<sup>a</sup> | 0.10 | 0.17 | 0.57 | 0.83 | -0.06 | 0.21 | 0.77 | 0.99 | 0.32 | 0.18 | 0.07 | 0.17 | 0.35 | 0.18 | 0.06 | 0.11 |
| General mean diffusivity<sup>a</sup> | 0.10 | 0.17 | 0.58 | 0.83 | 0.00 | 0.22 | 0.99 | 0.99 | 0.02 | 0.18 | 0.92 | 0.92 | 0.09 | 0.19 | 0.64 | 0.73 |
| *Mid- to late adulthood neighbourhood deprivation (n=359 to 396)* |  |  |  |  |  |  |  |  |  |  |  |  |  |  |  |  |
| Total brain volume | 0.01 | 0.06 | 0.90 | 0.97 | -0.09 | 0.07 | 0.22 | 0.52 | 0.05 | 0.07 | 0.50 | 0.72 | 0.10 | 0.07 | 0.12 | 0.31 |
| Grey matter volume | 0.09 | 0.09 | 0.31 | 0.82 | 0.08 | 0.11 | 0.46 | 0.52 | -0.01 | 0.10 | 0.89 | 0.89 | 0.25 | 0.10 | 0.01 | 0.08 |
| Normal-appearing white matter volume | -0.10 | 0.09 | 0.26 | 0.82 | -0.07 | 0.11 | 0.55 | 0.55 | 0.06 | 0.10 | 0.54 | 0.72 | -0.17 | 0.10 | 0.09 | 0.31 |
| White matter hyperintensity volume | 0.09 | 0.14 | 0.50 | 0.97 | -0.39 | 0.17 | 0.02 | 0.17 | 0.28 | 0.15 | 0.07 | 0.26 | 0.14 | 0.16 | 0.35 | 0.47 |
| Cortical surface area | 0.02 | 0.08 | 0.77 | 0.97 | -0.08 | 0.10 | 0.44 | 0.52 | 0.23 | 0.09 | 0.01 | 0.06 | 0.00 | 0.09 | 0.98 | 0.98 |
| Mean cortical thickness | 0.01 | 0.16 | 0.97 | 0.97 | -0.17 | 0.20 | 0.38 | 0.52 | -0.18 | 0.17 | 0.30 | 0.60 | 0.10 | 0.18 | 0.57 | 0.65 |
| General fractional anisotropy<sup>a</sup> | -0.06 | 0.17 | 0.71 | 0.97 | -0.20 | 0.20 | 0.32 | 0.52 | 0.22 | 0.18 | 0.22 | 0.60 | -0.24 | 0.19 | 0.19 | 0.39 |
| General mean diffusivity<sup>a</sup> | 0.20 | 0.17 | 0.25 | 0.82 | -0.18 | 0.21 | 0.41 | 0.52 | 0.06 | 0.19 | 0.73 | 0.84 | 0.21 | 0.19 | 0.28 | 0.44 |
| *Accumulated neighbourhood deprivation (n=255 to 281)* |  |  |  |  |  |  |  |  |  |  |  |  |  |  |  |  |
| Total brain volume | -0.04 | 0.07 | 0.52 | 0.93 | -0.10 | 0.09 | 0.30 | 0.57 | 0.15 | 0.08 | 0.04 | 0.17 | **0.21** | **0.07** | **0.003** | **0.01** |
| Grey matter volume | -0.01 | 0.10 | 0.96 | 0.96 | -0.11 | 0.15 | 0.43 | 0.57 | 0.12 | 0.12 | 0.30 | 0.50 | **0.31** | **0.11** | **0.003** | **0.01** |
| Normal-appearing white matter volume | -0.05 | 0.10 | 0.62 | 0.93 | 0.12 | 0.15 | 0.40 | 0.57 | 0.12 | 0.12 | 0.31 | 0.50 | -0.01 | 0.11 | 0.93 | 0.93 |
| White matter hyperintensity volume | 0.08 | 0.16 | 0.63 | 0.93 | -0.44 | 0.22 | 0.05 | 0.36 | 0.15 | 0.18 | 0.41 | 0.54 | 0.17 | 0.17 | 0.34 | 0.55 |
| Cortical surface area | 0.04 | 0.10 | 0.67 | 0.93 | -0.18 | 0.13 | 0.18 | 0.57 | 0.24 | 0.10 | 0.02 | 0.17 | 0.17 | 0.10 | 0.10 | 0.28 |
| Mean cortical thickness | 0.07 | 0.19 | 0.73 | 0.93 | -0.18 | 0.28 | 0.51 | 0.58 | -0.07 | 0.21 | 0.76 | 0.82 | 0.15 | 0.21 | 0.46 | 0.62 |
| General fractional anisotropy<sup>a</sup> | 0.23 | 0.20 | 0.25 | 0.93 | -0.04 | 0.27 | 0.89 | 0.89 | 0.40 | 0.21 | 0.06 | 0.17 | 0.30 | 0.21 | 0.16 | 0.33 |
| General mean diffusivity<sup>a</sup> | -0.05 | 0.21 | 0.82 | 0.93 | 0.27 | 0.29 | 0.36 | 0.57 | 0.05 | 0.23 | 0.82 | 0.82 | 0.10 | 0.23 | 0.65 | 0.77 |

Models with relevant interaction terms were fitted within the structural equation modelling framework applying full information maximum likelihood estimation. Total sample size was $N=658$ for total brain, grey matter and normal-appearing white matter volumes, $N=672$ for white matter hyperintensity volume, $N=636$ for cortical surface area and mean cortical thickness, and $N=665$ for general fractional anisotropy and mean diffusivity; this table indicates ranges of observations for each exposure-outcomes pairs on which reported effect sizes are based ( $n$ ). Bold typeface denotes false discovery rate adjusted significance ( $p_{FDR}$ ). SE = standard error.
Models were adjusted for sex, age, (intracranial volume for macrostructural measures,) father's occupational social class and APOE $\epsilon 4$ allele status. In addition, young adulthood models were adjusted for childhood IQ and years spent in education, and mid- to late adulthood/ accumulation models also for adult occupational social class.
<sup>a</sup> No adjustment for intracranial volume.

**Supplementary Table 8:**
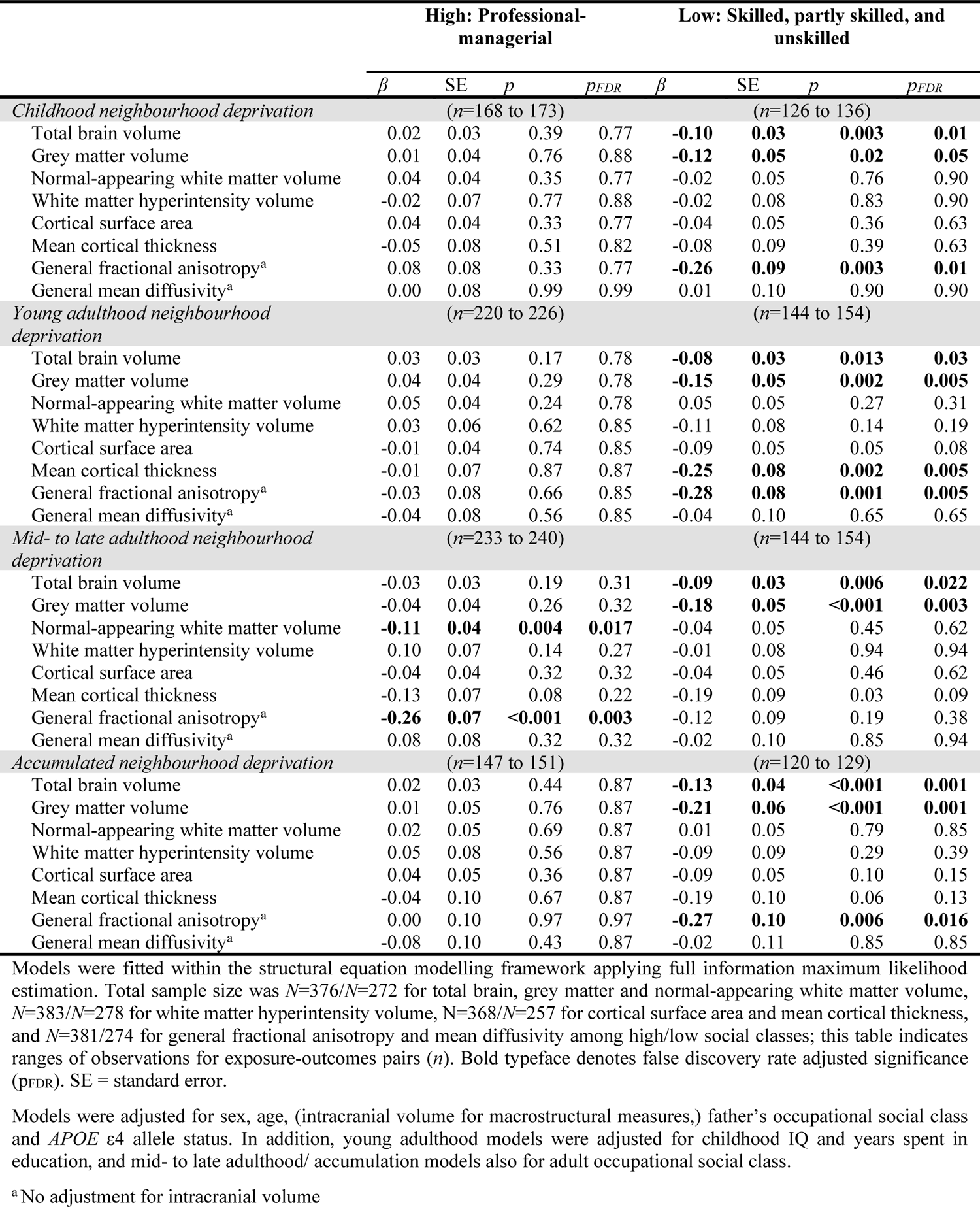
Association between life-course models of neighbourhood deprivation and global brain outcomes stratified by adult occupational social class

**Supplementary Table 9:** Association between life-course models of neighbourhood deprivation and global brain outcomes after considering exposure during previous epoch

| | $\beta$ | SE | $p$ | $p_{FDR}$ | % Change from Model 2<sup>a</sup> |
| --- | --- | --- | --- | --- | --- |
| *Young adulthood neighbourhood deprivation (n=367 to 383)* |  |  |  |  |  |
| Total brain volume | 0.01 | 0.02 | 0.69 | 0.78 | -150.00% |
| Grey matter volume | -0.03 | 0.04 | 0.47 | 0.63 | -40.00% |
| Normal-appearing white matter volume | 0.07 | 0.04 | 0.08 | 0.21 | 40.00% |
| White matter hyperintensity volume | -0.02 | 0.06 | 0.78 | 0.78 | -55.00% |
| Cortical surface area | -0.05 | 0.04 | 0.15 | 0.30 | 0.00% |
| Mean cortical thickness | -0.12 | 0.07 | 0.07 | 0.21 | -20.00% |
| General fractional anisotropy<sup>b</sup> | -0.13 | 0.07 | 0.06 | 0.21 | -7.14% |
| General mean diffusivity<sup>b</sup> | -0.09 | 0.07 | 0.22 | 0.35 | 50.00% |
| *Mid- to late adulthood neighbourhood deprivation (n=379 to 396)* |  |  |  |  |  |
| Total brain volume | **-0.05** | **0.02** | **0.03** | **0.04** | **-16.67%** |
| Grey matter volume | **-0.09** | **0.03** | **0.01** | **0.03** | **-18.18%** |
| Normal-appearing white matter volume | **-0.09** | **0.03** | **0.01** | **0.03** | **28.57%** |
| White matter hyperintensity volume | 0.07 | 0.05 | 0.22 | 0.29 | 40.00% |
| Cortical surface area | -0.01 | 0.03 | 0.67 | 0.76 | -75.00% |
| Mean cortical thickness | **-0.15** | **0.06** | **0.02** | **0.03** | **0.00%** |
| General fractional anisotropy<sup>b</sup> | **-0.17** | **0.06** | **0.01** | **0.03** | **-10.53%** |
| General mean diffusivity<sup>b</sup> | 0.01 | 0.07 | 0.86 | 0.86 | -66.67% |

Models were fitted within the structural equation modelling framework applying full information maximum likelihood estimation; exposure of interest was regressed on exposure during the previous period (i.e., young adulthood on childhood, mid- to late adulthood on young adulthood neighbourhood deprivation). Models for childhood neighbourhood deprivation and accumulated neighbourhood deprivation are not presented as this sensitivity analysis cannot be applied for them. Total sample size was $N=658$ for total brain, grey matter and normal-appearing white matter volumes, $N=672$ for white matter hyperintensity volume, $N=636$ for cortical surface area and mean cortical thickness, and $N=665$ for general fractional anisotropy and mean diffusivity; this table indicates ranges of observations for each exposure-outcomes pairs on which reported effect sizes are based ( $n$ ). Bold typeface denotes false discovery rate adjusted significance ( $p_{FDR}$ ). SE = standard error.
Models were adjusted for sex, age, (intracranial volume for macrostructural measures,) father's occupational social class, *APOE* $\epsilon 4$ allele status, childhood IQ, years spent in education and neighbourhood deprivation during previous exposure period, mid- to late adulthood models also adjusted for adult occupational social class.
<sup>a</sup> Main models did not include neighbourhood deprivation in previous epochs.
<sup>b</sup> No adjustment for intracranial volume.

**Supplementary Table 10:** Association between life-course models of neighbourhood deprivation and global brain outcomes after adjusting for stroke identified from MRI scans

|  | Model 2 – S2 |  |  |  |
| --- | --- | --- | --- | --- |
| | $\beta$ | SE | $p$ | $p_{FDR}$ |
| *Childhood neighbourhood deprivation (n=296 to 311)* |  |  |  |  |
| Total brain volume | -0.04 | 0.02 | 0.10 | 0.39 |
| Grey matter volume | -0.05 | 0.03 | 0.09 | 0.39 |
| Normal-appearing white matter volume | 0.02 | 0.03 | 0.58 | 0.77 |
| White matter hyperintensity volume | -0.03 | 0.05 | 0.57 | 0.77 |
| Cortical surface area | -0.01 | 0.03 | 0.84 | 0.96 |
| Mean cortical thickness | -0.07 | 0.06 | 0.23 | 0.61 |
| General fractional anisotropy<sup>a</sup> | -0.06 | 0.06 | 0.30 | 0.61 |
| General mean diffusivity<sup>a</sup> | 0.00 | 0.06 | 0.98 | 0.98 |
| *Young adulthood neighbourhood social deprivation (n=367 to 383)* |  |  |  |  |
| Total brain volume | -0.02 | 0.02 | 0.34 | 0.44 |
| Grey matter volume | -0.05 | 0.03 | 0.11 | 0.22 |
| Normal-appearing white matter volume | 0.05 | 0.03 | 0.14 | 0.22 |
| White matter hyperintensity volume | -0.03 | 0.05 | 0.48 | 0.48 |
| Cortical surface area | -0.05 | 0.03 | 0.06 | 0.17 |
| Mean cortical thickness | -0.10 | 0.05 | 0.06 | 0.17 |
| General fractional anisotropy<sup>a</sup> | -0.14 | 0.06 | 0.01 | 0.11 |
| General mean diffusivity<sup>a</sup> | -0.05 | 0.06 | 0.39 | 0.44 |
| *Mid- to late adulthood neighbourhood deprivation (n=379 to 396)* |  |  |  |  |
| Total brain volume | **-0.06** | **0.02** | **0.01** | **0.02** |
| Grey matter volume | **-0.10** | **0.03** | **0.002** | **0.009** |
| Normal-appearing white matter volume | -0.06 | 0.03 | 0.07 | 0.11 |
| White matter hyperintensity volume | 0.04 | 0.05 | 0.46 | 0.52 |
| Cortical surface area | -0.04 | 0.03 | 0.16 | 0.21 |
| Mean cortical thickness | **-0.15** | **0.06** | **0.01** | **0.02** |
| General fractional anisotropy<sup>a</sup> | **-0.18** | **0.06** | **0.002** | **0.009** |
| General mean diffusivity<sup>a</sup> | 0.01 | 0.06 | 0.83 | 0.83 |
| *Accumulated neighbourhood deprivation (n=268 to 281)* |  |  |  |  |
| Total brain volume | -0.05 | 0.03 | 0.05 | 0.20 |
| Grey matter volume | -0.09 | 0.04 | 0.02 | 0.19 |
| Normal-appearing white matter volume | 0.02 | 0.04 | 0.53 | 0.57 |
| White matter hyperintensity volume | -0.03 | 0.06 | 0.57 | 0.57 |
| Cortical surface area | -0.02 | 0.04 | 0.53 | 0.57 |
| Mean cortical thickness | -0.11 | 0.07 | 0.15 | 0.29 |
| General fractional anisotropy<sup>a</sup> | -0.13 | 0.07 | 0.08 | 0.22 |
| General mean diffusivity<sup>a</sup> | -0.07 | 0.08 | 0.38 | 0.57 |

Models were fitted within the structural equation modelling framework applying full information maximum likelihood estimation. Total sample size was N=658 for total brain, grey matter and normal-appearing white matter volume, N=672 for white matter hyperintensity volume, N=636 for cortical surface area and mean cortical thickness, and N=665 for general fractional anisotropy and mean diffusivity; this table indicates ranges of observations for each exposure-outcomes pairs on which reported effect sizes are based (n). Bold typeface denotes false discovery rate adjusted significance ( $p_{FDR}$ ). SE = standard error.
Models were adjusted for sex, age, MRI-based stroke, (intracranial volume for macrostructural measures,) father's occupational social class and *APOE* $\epsilon 4$ allele status. In addition, young adulthood models were adjusted for childhood IQ and years spent in education, and mid- to late adulthood / accumulation models also for adult occupational social class.
<sup>a</sup> No adjustment for intracranial volume

**Supplementary Table 11:** Association between mid-to-late adulthood neighbourhood deprivation and global brain outcomes after adjusting for late adulthood health status

|  | Model 2 – S3 |  |  |  | % Change<br>from Model 2<sup>a</sup> |
| --- | --- | --- | --- | --- | --- |
| | $\beta$ | SE | $p$ | $p_{FDR}$ | |
| *Mid- to late adulthood neighbourhood deprivation (n=379 to 396)* |  |  |  |  |  |
| Total brain volume | -0.05 | 0.02 | 0.02 | 0.05 | -16.67% |
| Grey matter volume | **-0.09** | **0.03** | **0.004** | **0.02** | **-18.18%** |
| Normal-appearing white matter volume | -0.05 | 0.03 | 0.10 | 0.15 | -28.57% |
| White matter hyperintensity volume | 0.04 | 0.05 | 0.42 | 0.48 | -20.00% |
| Cortical surface area | -0.04 | 0.03 | 0.20 | 0.27 | 0.00% |
| Mean cortical thickness | -0.10 | 0.06 | 0.07 | 0.15 | -33.33% |
| General fractional anisotropy<sup>b</sup> | **-0.17** | **0.06** | **0.004** | **0.02** | **-10.53%** |
| General mean diffusivity<sup>b</sup> | 0.02 | 0.06 | 0.71 | 0.71 | -33.33% |

Models were fitted within the structural equation modelling framework applying full information maximum likelihood estimation. Total sample size was $N=658$ for total brain, grey matter and normal-appearing white matter volumes, $N=672$ for white matter hyperintensity volume, $N=636$ for cortical surface area and mean cortical thickness, and $N=665$ for general fractional anisotropy and mean diffusivity; this table indicates ranges of observations for each exposure-outcomes pairs on which reported effect sizes are based ( $n$ ). Bold typeface denotes false discovery rate adjusted significance ( $p_{FDR}$ ). SE = standard error.
Models were adjusted for sex, age, (intracranial volume for volumetric measures,) father's occupational social class, *APOE* $\epsilon 4$ allele status, childhood IQ, years spent in education, adult occupational social class, smoking status, BMI, and history of hypertension, diabetes, stroke, and cardiovascular diseases.
<sup>a</sup> Main models did not include health-related variables.
<sup>b</sup> No adjustment for intracranial volume

**Supplementary Table 12:**
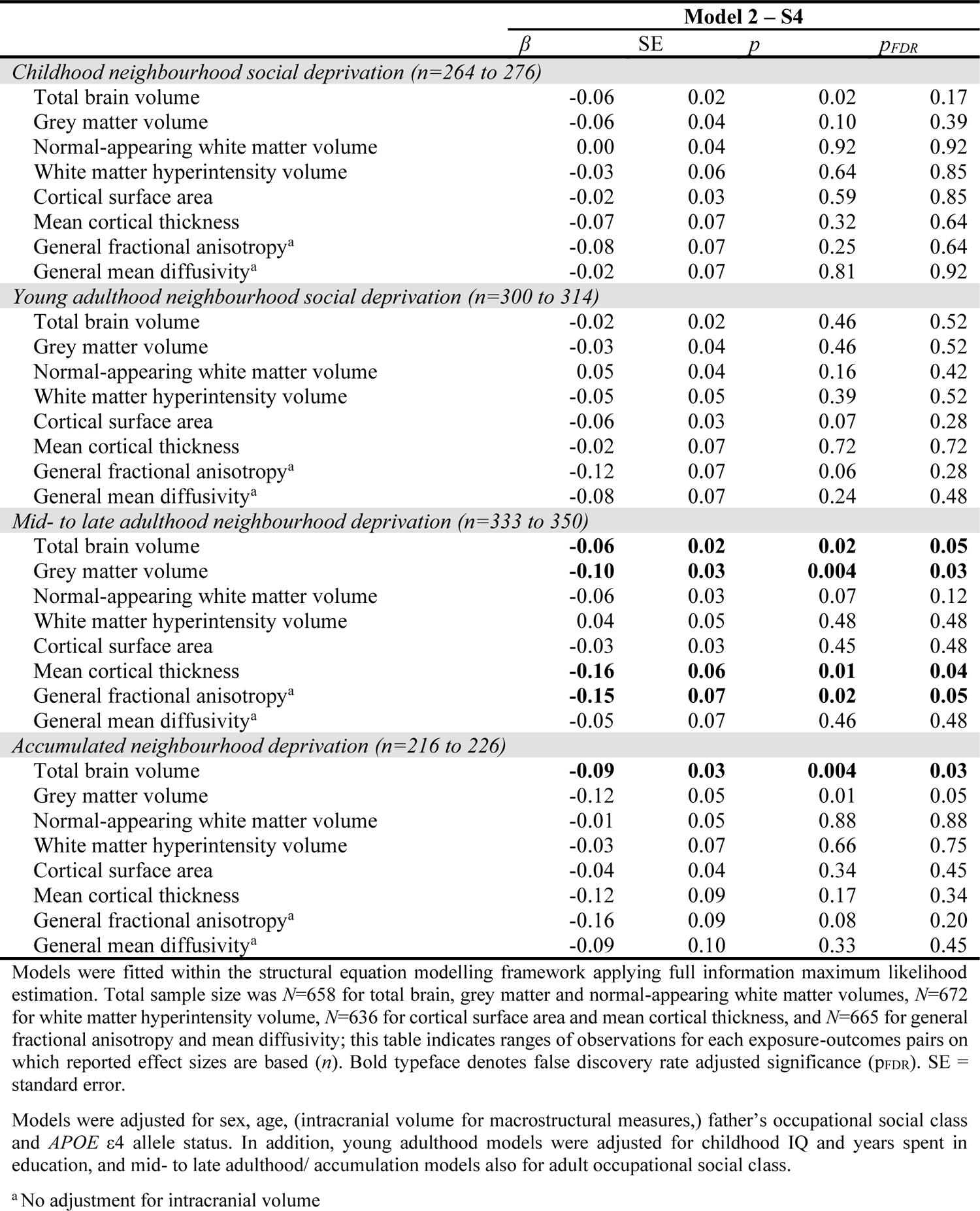
Association between life-course models of neighbourhood deprivation and global brain outcomes among those living in Edinburgh throughout each decade of the exposure periods

|  | **Model 2 – S4** |  |  |  |
| --- | --- | --- | --- | --- |
| | $\beta$ | SE | $p$ | $p_{FDR}$ |
| *Childhood neighbourhood social deprivation (n=264 to 276)* |  |  |  |  |
| Total brain volume | -0.06 | 0.02 | 0.02 | 0.17 |
| Grey matter volume | -0.06 | 0.04 | 0.10 | 0.39 |
| Normal-appearing white matter volume | 0.00 | 0.04 | 0.92 | 0.92 |
| White matter hyperintensity volume | -0.03 | 0.06 | 0.64 | 0.85 |
| Cortical surface area | -0.02 | 0.03 | 0.59 | 0.85 |
| Mean cortical thickness | -0.07 | 0.07 | 0.32 | 0.64 |
| General fractional anisotropy<sup>a</sup> | -0.08 | 0.07 | 0.25 | 0.64 |
| General mean diffusivity<sup>a</sup> | -0.02 | 0.07 | 0.81 | 0.92 |
| *Young adulthood neighbourhood social deprivation (n=300 to 314)* |  |  |  |  |
| Total brain volume | -0.02 | 0.02 | 0.46 | 0.52 |
| Grey matter volume | -0.03 | 0.04 | 0.46 | 0.52 |
| Normal-appearing white matter volume | 0.05 | 0.04 | 0.16 | 0.42 |
| White matter hyperintensity volume | -0.05 | 0.05 | 0.39 | 0.52 |
| Cortical surface area | -0.06 | 0.03 | 0.07 | 0.28 |
| Mean cortical thickness | -0.02 | 0.07 | 0.72 | 0.72 |
| General fractional anisotropy<sup>a</sup> | -0.12 | 0.07 | 0.06 | 0.28 |
| General mean diffusivity<sup>a</sup> | -0.08 | 0.07 | 0.24 | 0.48 |
| *Mid- to late adulthood neighbourhood deprivation (n=333 to 350)* |  |  |  |  |
| Total brain volume | **-0.06** | **0.02** | **0.02** | **0.05** |
| Grey matter volume | **-0.10** | **0.03** | **0.004** | **0.03** |
| Normal-appearing white matter volume | -0.06 | 0.03 | 0.07 | 0.12 |
| White matter hyperintensity volume | 0.04 | 0.05 | 0.48 | 0.48 |
| Cortical surface area | -0.03 | 0.03 | 0.45 | 0.48 |
| Mean cortical thickness | **-0.16** | **0.06** | **0.01** | **0.04** |
| General fractional anisotropy<sup>a</sup> | **-0.15** | **0.07** | **0.02** | **0.05** |
| General mean diffusivity<sup>a</sup> | -0.05 | 0.07 | 0.46 | 0.48 |
| *Accumulated neighbourhood deprivation (n=216 to 226)* |  |  |  |  |
| Total brain volume | **-0.09** | **0.03** | **0.004** | **0.03** |
| Grey matter volume | -0.12 | 0.05 | 0.01 | 0.05 |
| Normal-appearing white matter volume | -0.01 | 0.05 | 0.88 | 0.88 |
| White matter hyperintensity volume | -0.03 | 0.07 | 0.66 | 0.75 |
| Cortical surface area | -0.04 | 0.04 | 0.34 | 0.45 |
| Mean cortical thickness | -0.12 | 0.09 | 0.17 | 0.34 |
| General fractional anisotropy<sup>a</sup> | -0.16 | 0.09 | 0.08 | 0.20 |
| General mean diffusivity<sup>a</sup> | -0.09 | 0.10 | 0.33 | 0.45 |

Models were fitted within the structural equation modelling framework applying full information maximum likelihood estimation. Total sample size was $N=658$ for total brain, grey matter and normal-appearing white matter volumes, $N=672$ for white matter hyperintensity volume, $N=636$ for cortical surface area and mean cortical thickness, and $N=665$ for general fractional anisotropy and mean diffusivity; this table indicates ranges of observations for each exposure-outcomes pairs on which reported effect sizes are based ( $n$ ). Bold typeface denotes false discovery rate adjusted significance ( $p_{FDR}$ ). SE = standard error.
Models were adjusted for sex, age, (intracranial volume for macrostructural measures,) father's occupational social class and *APOE* $\epsilon 4$ allele status. In addition, young adulthood models were adjusted for childhood IQ and years spent in education, and mid- to late adulthood/ accumulation models also for adult occupational social class.
<sup>a</sup> No adjustment for intracranial volume

**Supplementary Table 13:** Association between life-course models of neighbourhood deprivation and global brain outcomes among individuals without cognitive impairment

|  | Model 2 – S5 |  |  |  |
| --- | --- | --- | --- | --- |
| | $\beta$ | SE | $p$ | $p_{FDR}$ |
| *Childhood neighbourhood social deprivation (n=246 to 255)* |  |  |  |  |
| Total brain volume | -0.05 | 0.02 | 0.01 | 0.10 |
| Grey matter volume | -0.05 | 0.03 | 0.11 | 0.44 |
| Normal-appearing white matter volume | 0.00 | 0.03 | 1.00 | 1.00 |
| White matter hyperintensity volume | -0.05 | 0.05 | 0.34 | 0.54 |
| Cortical surface area | -0.01 | 0.03 | 0.70 | 0.80 |
| Mean cortical thickness | -0.08 | 0.06 | 0.20 | 0.53 |
| General fractional anisotropy<sup>a</sup> | -0.07 | 0.07 | 0.30 | 0.54 |
| General mean diffusivity<sup>a</sup> | -0.03 | 0.07 | 0.61 | 0.80 |
| *Young adulthood neighbourhood social deprivation (n=280 to 292)* |  |  |  |  |
| Total brain volume | -0.03 | 0.02 | 0.22 | 0.28 |
| Grey matter volume | -0.04 | 0.03 | 0.21 | 0.28 |
| Normal-appearing white matter volume | 0.04 | 0.03 | 0.24 | 0.28 |
| White matter hyperintensity volume | -0.05 | 0.05 | 0.31 | 0.31 |
| Cortical surface area | -0.06 | 0.03 | 0.04 | 0.15 |
| Mean cortical thickness | -0.10 | 0.06 | 0.06 | 0.17 |
| General fractional anisotropy<sup>a</sup> | **-0.17** | **0.06** | **0.005** | **0.04** |
| General mean diffusivity<sup>a</sup> | -0.09 | 0.06 | 0.17 | 0.28 |
| *Mid- to late adulthood neighbourhood deprivation (n=313 to 328)* |  |  |  |  |
| Total brain volume | **-0.07** | **0.02** | **0.003** | **0.01** |
| Grey matter volume | **-0.11** | **0.03** | **0.001** | **0.008** |
| Normal-appearing white matter volume | **-0.09** | **0.03** | **0.006** | **0.01** |
| White matter hyperintensity volume | 0.05 | 0.05 | 0.39 | 0.44 |
| Cortical surface area | -0.04 | 0.03 | 0.18 | 0.23 |
| Mean cortical thickness | **-0.15** | **0.06** | **0.009** | **0.01** |
| General fractional anisotropy<sup>a</sup> | **-0.18** | **0.06** | **0.004** | **0.01** |
| General mean diffusivity<sup>a</sup> | 0.03 | 0.07 | 0.69 | 0.69 |
| *Accumulated neighbourhood deprivation (n=201 to 209)* |  |  |  |  |
| Total brain volume | **-0.07** | **0.03** | **0.006** | **0.047** |
| Grey matter volume | -0.09 | 0.04 | 0.02 | 0.10 |
| Normal-appearing white matter volume | -0.02 | 0.04 | 0.61 | 0.61 |
| White matter hyperintensity volume | -0.04 | 0.06 | 0.50 | 0.58 |
| Cortical surface area | -0.03 | 0.04 | 0.37 | 0.51 |
| Mean cortical thickness | -0.12 | 0.08 | 0.13 | 0.26 |
| General fractional anisotropy<sup>a</sup> | -0.15 | 0.08 | 0.06 | 0.15 |
| General mean diffusivity<sup>a</sup> | -0.07 | 0.08 | 0.39 | 0.51 |

Models were fitted within the structural equation modelling framework applying full information maximum likelihood estimation. Cognitive impairment was defined as either having a diagnosis of dementia or scoring <24 points in the Mini Mental State Examination. Total sample size was $N=611$ for total brain, grey matter and normal-appearing white matter volume, $N=625$ for white matter hyperintensity volume, $N=594$ for cortical surface area and mean cortical thickness, and $N=620$ for general fractional anisotropy and mean diffusivity; ranges of observations for exposure-outcomes pairs are presented in the table ( $n$ ). Bold typeface denotes false discovery rate adjusted significance ( $p_{FDR}$ ). SE = standard error.
Models were adjusted for sex, age, (intracranial volume for macrostructural measures,) father's occupational social class and *APOE* $\epsilon 4$ allele status. In addition, young adulthood models were adjusted for childhood IQ and years spent in education, and mid- to late adulthood/ accumulation models also for adult occupational social class.
<sup>a</sup> No adjustment for intracranial volume

**Supplementary Table 14:** Complete case analysis testing the association between life-course models of neighbourhood deprivation and global brain outcomes

|  | Model 2 – S6 |  |  |  |
| --- | --- | --- | --- | --- |
| | $\beta$ | SE | $p$ | $p_{FDR}$ |
| *Childhood neighbourhood deprivation* |  |  |  |  |
| Total brain volume ($N=261$) | -0.05 | 0.02 | 0.02 | 0.13 |
| Grey matter volume ($N=261$) | -0.05 | 0.04 | 0.16 | 0.54 |
| Normal-appearing white matter volume ($N=261$) | 0.00 | 0.03 | 0.94 | 0.94 |
| White matter hyperintensity volume ($N=266$) | -0.06 | 0.07 | 0.40 | 0.64 |
| Cortical surface area ($N=255$) | -0.02 | 0.04 | 0.64 | 0.86 |
| Mean cortical thickness ($N=255$) | -0.07 | 0.07 | 0.27 | 0.54 |
| General fractional anisotropy<sup>a</sup> ($N=263$) | -0.08 | 0.07 | 0.25 | 0.54 |
| General mean diffusivity<sup>a</sup> ($N=263$) | -0.02 | 0.06 | 0.81 | 0.92 |
| *Young adulthood neighbourhood deprivation* |  |  |  |  |
| Total brain volume ($N=310$) | -0.02 | 0.02 | 0.27 | 0.44 |
| Grey matter volume ($N=310$) | -0.03 | 0.03 | 0.39 | 0.46 |
| Normal-appearing white matter volume ($N=310$) | 0.02 | 0.03 | 0.46 | 0.46 |
| White matter hyperintensity volume ($N=316$) | -0.07 | 0.07 | 0.28 | 0.44 |
| Cortical surface area ($N=304$) | -0.08 | 0.03 | 0.02 | 0.09 |
| Mean cortical thickness ($N=304$) | -0.09 | 0.06 | 0.17 | 0.44 |
| General fractional anisotropy<sup>a</sup> ($N=311$) | **-0.18** | **0.06** | **0.005** | **0.04** |
| General mean diffusivity<sup>a</sup> ($N=311$) | -0.05 | 0.06 | 0.41 | 0.46 |
| *Mid- to late adulthood neighbourhood deprivation* |  |  |  |  |
| Total brain volume ($N=321$) | -0.05 | 0.02 | 0.03 | 0.07 |
| Grey matter volume ($N=321$) | -0.07 | 0.03 | 0.04 | 0.08 |
| Normal-appearing white matter volume ($N=321$) | -0.07 | 0.03 | 0.01 | 0.06 |
| White matter hyperintensity volume ($N=326$) | 0.03 | 0.07 | 0.67 | 0.68 |
| Cortical surface area ($N=313$) | -0.04 | 0.04 | 0.33 | 0.45 |
| Mean cortical thickness ($N=313$) | -0.13 | 0.07 | 0.06 | 0.09 |
| General fractional anisotropy<sup>a</sup> ($N=321$) | **-0.20** | **0.06** | **0.002** | **0.02** |
| General mean diffusivity<sup>a</sup> ($N=321$) | 0.03 | 0.06 | 0.68 | 0.68 |
| *Accumulated neighbourhood deprivation* |  |  |  |  |
| Total brain volume ($N=228$) | -0.06 | 0.03 | 0.02 | 0.18 |
| Grey matter volume ($N=228$) | -0.06 | 0.04 | 0.11 | 0.29 |
| Normal-appearing white matter volume ($N=228$) | -0.01 | 0.04 | 0.70 | 0.70 |
| White matter hyperintensity volume ($N=232$) | -0.06 | 0.09 | 0.52 | 0.60 |
| Cortical surface area ($N=222$) | -0.04 | 0.04 | 0.36 | 0.49 |
| Mean cortical thickness ($N=222$) | -0.07 | 0.08 | 0.37 | 0.49 |
| General fractional anisotropy<sup>a</sup> ($N=229$) | -0.15 | 0.08 | 0.06 | 0.23 |
| General mean diffusivity<sup>a</sup> ($N=229$) | -0.07 | 0.06 | 0.30 | 0.49 |

Models were fitted with multivariate linear regression applying complete case analysis; therefore, underlying sample sizes differ across the presented estimates. Latent factors of general fractional anisotropy and general mean diffusivity were predicted and extracted from the measurement model. Sample sizes for each model are indicated in the table ( $N$ ). Bold typeface denotes false discovery rate adjusted significance ( $p_{FDR}$ ). SE = standard error.
Models were adjusted for sex, age, (intracranial volume for macrostructural measures,) father's occupational social class and *APOE* $\epsilon 4$ allele status. In addition, young adulthood models were adjusted for childhood IQ and years spent in education, and mid- to late adulthood/ accumulation models also for adult occupational social class.
<sup>a</sup> No adjustment for intracranial volume

**Supplementary Figure 8:**
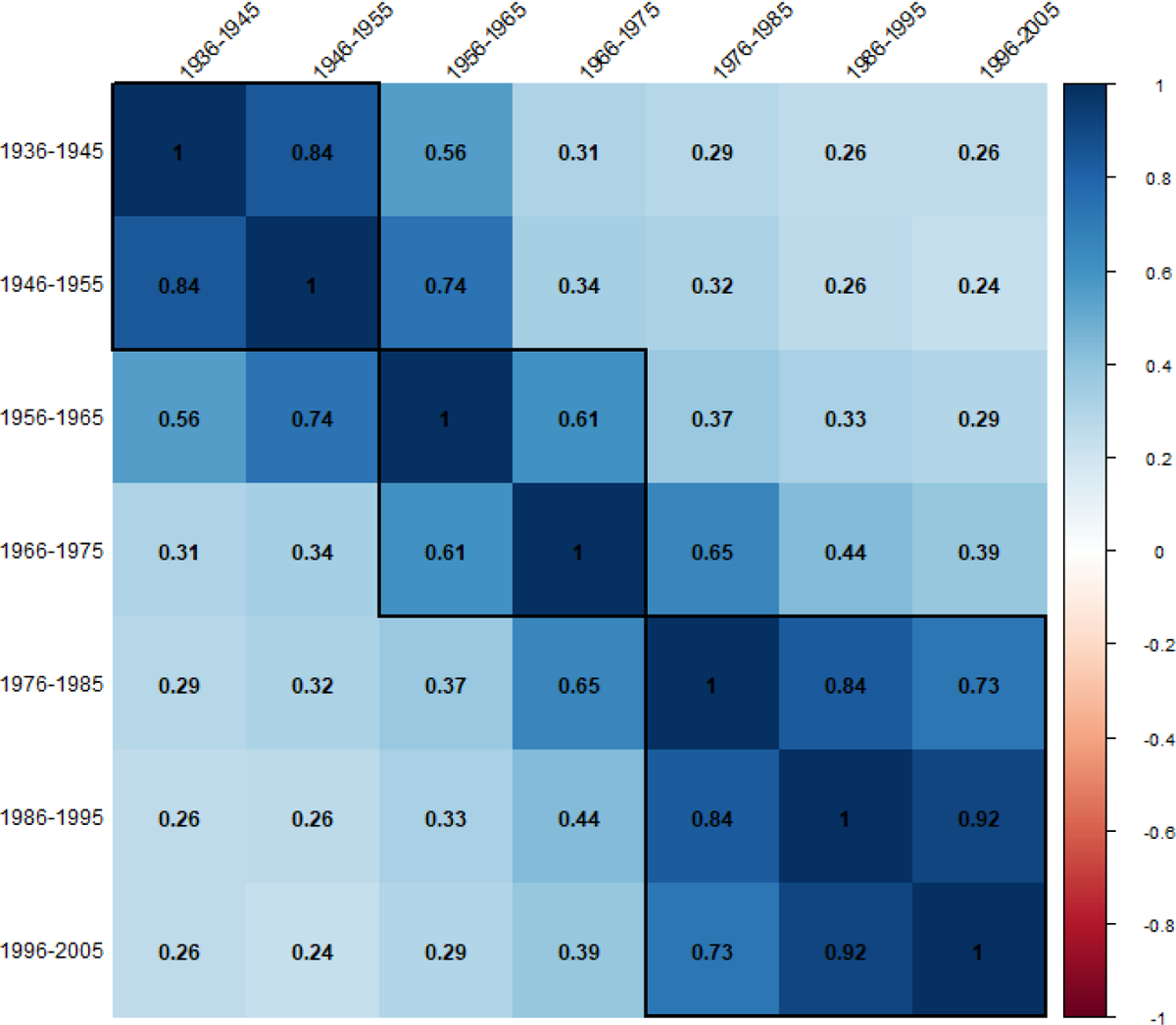
Pearson’s correlation coefficients between individual exposures to neighbourhood deprivation. Pairwise deletion was applied for missing values to preserve the maximum amount of information. Black rectangles indicate childhood (1936-1955), young adulthood (1956-1975), and mid- to late adulthood (1976-2005) periods. All presented correlation coefficients were significant (*p* < 0.001).

**Supplementary Figure 9:**
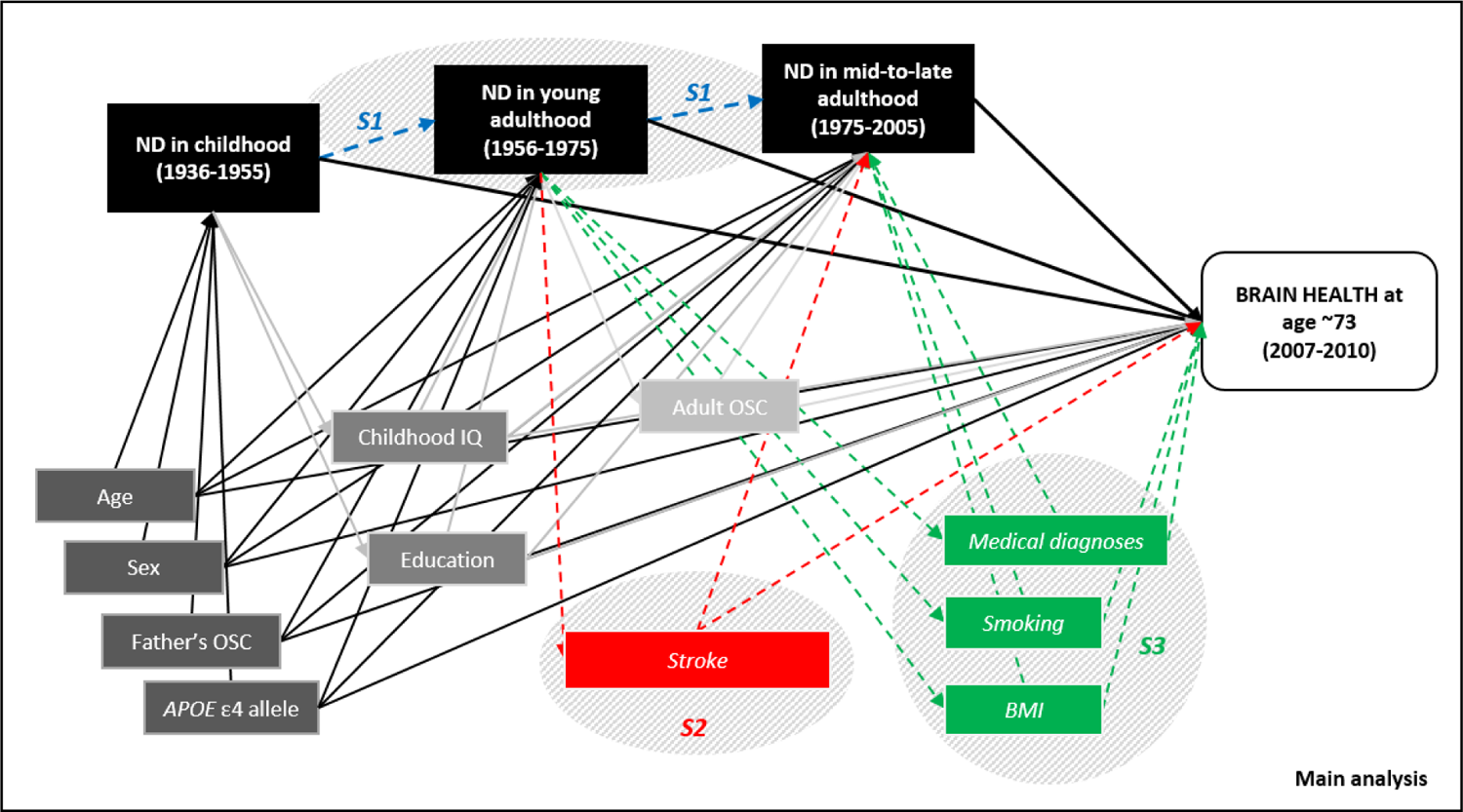
Directed acyclic graph representing associations between neighbourhood deprivation, global brain outcomes and confounders considered in the main and sensitivity analyses. Confounders included in the main analyses are coloured grey: dark grey are considered as confounders for all life course models, medium grey for young adulthood and mid- to late adulthood exposures, light grey for mid- to late adulthood exposures only. Sensitivity analyses considering selective mobility (S1) and potential health-related confounders (S2, S3) are blue, red, and green, respectively. Links between confounders are not shown for simplicity. BMI = body mass index; IQ = intelligence quotient; ND = neighbourhood deprivation; OSC = occupational social class.

**Supplementary Table 15:** Standardized tract loadings, explained variance and goodness of fit indices for fractional anisotropy and mean diffusivity

|  | Fractional anisotropy | Mean diffusivity |
| --- | --- | --- |
| *Standardized tract loadings* |  |  |
| Genu of corpus callosum | 0.57 | 0.57 |
| Splenium of corpus callosum | 0.41 | 0.28 |
| Left arcuate fasciculus | 0.65 | 0.70 |
| Right arcuate fasciculus | 0.65 | 0.74 |
| Left anterior thalamic radiation | 0.62 | 0.67 |
| Right anterior thalamic radiation | 0.60 | 0.58 |
| Left rostral cingulum | 0.52 | 0.60 |
| Right rostral cingulum | 0.53 | 0.72 |
| Left inferior longitudinal fasciculus | 0.46 | 0.37 |
| Right inferior longitudinal fasciculus | 0.44 | 0.36 |
| Left uncinate fasciculus | 0.62 | 0.63 |
| Right uncinate fasciculus | 0.65 | 0.68 |
| *% of variance explained* |  |  |
|  | 32.0% | 35.2% |
| *Goodness of fit indices* |  |  |
| Comparative Fit Index | 0.97 | 0.97 |
| Tucker Lewis Index | 0.96 | 0.96 |
| Root Mean Square Error of Approximation | 0.04 | 0.05 |
| Standardised Root Mean Square Residual | 0.03 | 0.03 |
| Chi-squared<sub>(48)</sub> | 106.34 | 131.43 |
| *p*-value | <0.001 | <0.001 |

Reference values are: Comparative Fit Index>0.95; Tucker-Lewis Index>0.95; Root Mean Square Error of Approximation<0.06; and Standardized Root Mean Square Residual<0.08 (1).

